# *Andes* Hantavirus human-to-human transmission dynamics and preparedness scenarios

**DOI:** 10.64898/2026.08.31.26361797

**Authors:** Daniele Proverbio, Giulia Giordano

## Abstract

Human-to-human transmission of the *Andes* virus (ANDV), which causes the deadly Hantavirus Pulmonary Syndrome, has been documented in several outbreaks, but remains insufficiently characterized to ensure preparedness against future emergence. Here, we integrate epidemiological data from historical outbreaks and from the 2026 *MV Hondius* outbreak with a stochastic network-based transmission model to quantify ANDV epidemic potential and control requirements. We estimate a reproduction number of *R*0 = 2.30 (95% CI: 1.33–3.27), supporting sustained human transmission. Uncertainty and sensitivity analysis show that variation in transmissibility and incubation period mainly affects outbreak timing and size, while superspreading events can substantially alter the epidemic evolution. Model analysis and simulations further reveal how contact-network structure strongly modulates outbreak size, with close-contact clustering facilitating containment. Due to the virus characteristics, rapid identification and isolation can effectively suppress outbreaks, even more effectively than in the case of COVID-19, whereas pre-symptomatic transmission or increased mixing could substantially increase risk. Our results provide a quantitative framework for ANDV preparedness and highlight the most critical transmission features for future surveillance.

## 1 Introduction

The disease outbreak on the cruise ship *MV Hondius* during spring 2026 was confirmed to be caused by *Andes* virus (ANDV), with 13 confirmed or probable cases and 3 deaths. It is the latest of several ANDV outbreaks reported over the past 30 years, which attests to the continuing public health threat posed by this group of viruses [1]. ANDV causes the highly lethal Hantavirus Pulmonary Syndrome (HPS) [2, 3, 4] and is the only hantavirus hypothesised to transmit between humans [5].

Most hantaviruses are transmitted exclusively through inhalation of aerosolized rodent secreta or excreta and remain strictly zoonotic [6, 7]. Consequently, surveillance and modelling have focused on rodent populations, rodent-to-rodent transmission, and spillover to humans [8, 9, 10, 11, 12]. In contrast, repeated evidence of human-to-human transmission has been reported for ANDV [13, 14, 15, 16, 17, 18, 19] and is hypothesised to explain the *MV Hondius* outbreak. Although the transmission mechanism remains uncertain, with saliva, aerosols and other routes being under investigation [20, 21], the possibility of sustained horizontal transmission calls for preventive modelling and preparedness [22, 23]. Integrating epidemiological evidence into mechanistic models is therefore essential for anticipating future outbreaks, particularly if ANDV evolves into more transmissible variants.

Mechanistic modelling of human-to-human ANDV transmission remain limited [24, 25] and the development, parametrization and analysis of predictive models for the *Andes* hantavirus are greatly missing. Current evidence indicates a long incubation period and transmission primarily through prolonged close contact with symptomatic individuals. These factors have so far enabled effective contact tracing and outbreak containment [13]. However, mutations that increase transmissibility or reduce detectability could substantially elevate epidemic risk given the virus’s high fatality rate. Beyond medical management and quarantine protocols [1, 26], predictive models are needed [23] to characterize current transmission dynamics, assess plausible outbreak scenarios, and support rapid adaptation of response strategies.

Here, we develop outbreak scenarios for ANDV based on empirical evidence. We first integrate epidemiological data from past and recent outbreaks to estimate key parameters, including the infectious period and the reproduction number *R*_0_. A meta-analysis across documented outbreaks strengthens the evidence for human-to-human transmission and reduces uncertainty in estimates of *R*_0_. We then develop a network-based transmission model to quantify the impact of parameter uncertainty on outbreak predictions, explain the spread aboard the *MV Hondius*, and estimate the probability of secondary outbreaks. We observe that, for ANDV, the structure of contacts can be as important as the biological transmissibility of the virus. Finally, we evaluate plausible quantitative scenarios involving different contact network structures, superspreading events [13] and pre-symptomatic infectiousness [21], and assess the effectiveness of simple mitigation measures.

Overall, this work provides an integrative, model-based assessment of human-to-human ANDV transmission, improves understanding of past outbreaks, and supports preparedness by identifying the transmission mechanisms and epidemiological conditions that can most likely increase the risk of future outbreaks.

## 2 Results

We first summarize the key events of the Spring 2026 ANDV outbreak, reconstructed from health agency reports and media sources. In April 2026, an HPS outbreak occurred aboard the luxury cruise ship *MV Hondius* [27]. The ship departed from Ushuaia (Argentina) on April 1st with 114 passengers and 61 crew members, according to the 2026/05/08 *Oceanwide Expeditions* press update (https://oceanwide-expeditions.com/press/press-update-m-v-hondius-8-may-2026-19-00-hrs-cet).

The first patient (P1) developed symptoms on April 6th and died on April 11th. On April 24th, two additional individuals became symptomatic, including P1’s wife, who died on April 26th. On that day, several passengers disembarked at Saint Helena, leaving 149 people on board. A crew member developed symptoms on April 27th, followed by two more cases on April 28th, one of whom died on May 2nd. The ship’s doctor became symptomatic on April 30th, followed by another passenger on May 1st. On May 4th, laboratory testing confirmed *Andes* virus infection in at least one patient [28]; at the time of writing, all patients except P1 had been classified as confirmed cases. Since early May, passengers were instructed to reduce close contacts, wear masks, and frequently use hand sanitizer (Reuters, “Hantavirus-hit cruise ship heads to Spain after three people evacuated”, 2026/05/07). Between May 3rd and 6th, the ship anchored at Cabo Verde, where symptomatic patients disembarked, before continuing to the Canary Islands, where it arrived on May 10th. Passengers were then repatriated; additional positive cases were identified on May 11th (two cases), 12th, and 14th. All repatriated passengers entered 42 to 45 days of quarantine [26]. Two further cases were detected after repatriation, in the Netherlands on May 22nd and in Spain on May 25th, the latter in a previously quarantined passenger. The epidemic progression is shown in Fig. 1 (green triangles). The outbreak was declared over by the WHO on July 2nd (BBC, “WHO says Hantavirus outbreak linked to ship is over”, 2026/07/02). Overall, 12 confirmed and 1 probable cases, including 3 deaths, were reported [27]. The exact transmission chains remain unavailable at the time of writing.

**Figure 1:**
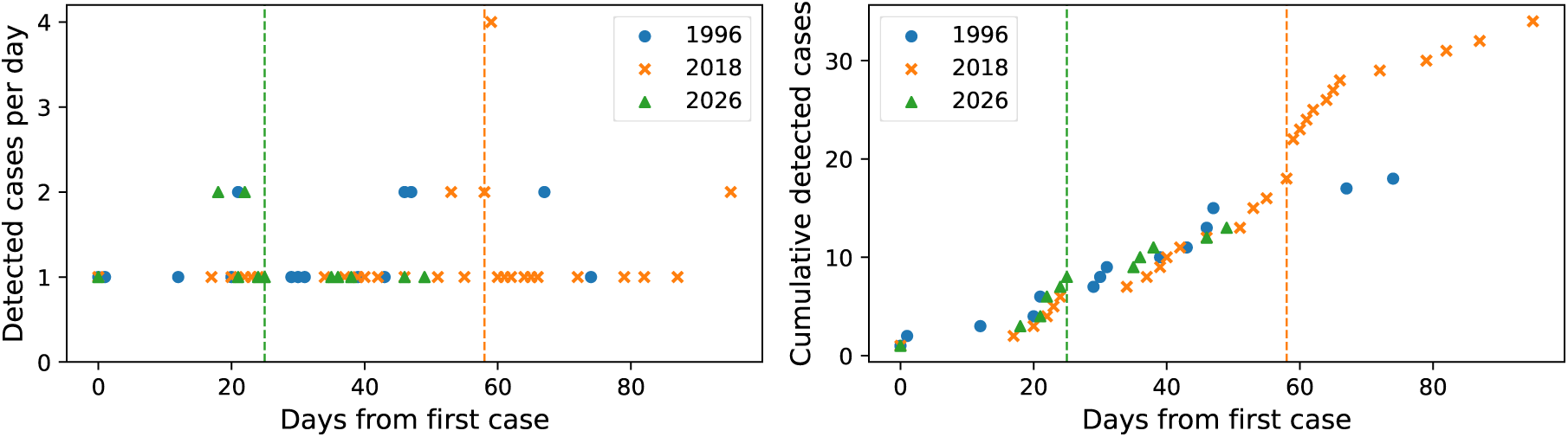
Left: reported incidence (detected cases per day) for the three main HPS outbreaks. Right: cumulative number of detected cases. Time is normalised from the day of detection of the first case (day 0): 22 September 1996, 3 November 2018 and 6 April 2026, respectively. The dashed lines indicate when control or protective measures began: the date is exact for the 2018 outbreak, approximated based on news reports for the 2026 outbreak, and unknown for the 1996 outbreak.

The 2026 outbreak adds to the record of relatively large and prolonged ANDV outbreaks described in the scientific literature. The first occurred between September and December 1996 in the towns of El Bolśon, Bariloche, and Esquel in southern Argentina, affecting residents, visitors, and several close contacts [15]. It provided the first evidence of person-to-person transmission of the virus. A total of 18 symptomatic cases were reported (Fig. 1, blue dots), including 9 deaths. A second major outbreak, associated with a superspreading event (see Supplementary Sec. S1) occurred between November 2018 and January 2019 in the village of Epuýen, Argentina, a few kilometres from the main area affected in 1996. Martínez et al. [13] documented 34 infections (Fig. 1, orange squares), resulting in 11 deaths. Details of these three outbreaks, the containment measures implemented and the associated datasets are provided in Methods.

### 2.1 Epidemiological evidence

The 1996, 2018, and 2026 outbreaks show similar temporal patterns of detected and cumulative cases, as illustrated in Fig. 1. In all three events, cases emerged gradually, with cumulative incidence increasing in a comparable manner. This suggests the hypothesis, later supported statistically, that the epidemiological characteristics of ANDV, including transmission probability and infectious period, have remained largely unchanged over the past 30 years. Figure 1 also provides qualitative insights into the effectiveness of control measures, whose implementation dates are indicated by dashed lines for the 2018 and 2026 outbreaks. The precise timing of control interventions in 1996 remains uncertain and is not reported in the figure. The 1996 and 2018 outbreaks were eventually contained through contact tracing and isolation of infected individuals. In 1996, transmission ceased approximately two months after isolating symptomatic patients and their close contacts; the few late cases were healthcare workers exposed to hospitalized patients and an individual infected in a car after a funeral. In 2018, it took about three months to suppress the outbreak, for which tracing and isolation were required. In the 2026 outbreak, isolation and treatment of cruise ship passengers were followed by only a small number of additional cases, likely representing infections acquired on board and detected later due to the virus’s long incubation period. In both the 2018 and 2026 outbreaks, approximately 30 days elapsed between the start of control measures and the identification of the final case, supporting the 42 to 45-day quarantine period recommended by WHO guidelines [26].

The Case Fatality Ratio (CFR) of HPS is relatively high. Based on cumulative evidence from past outbreaks and isolated cases, literature and public health sources estimate HPS CFR between 30% and 50%, with peaks up to 70% [1, 2]. CFRs during the three major outbreaks were consistently high: 50% in 1996 (9/18 cases) [15], 32% in 2018 (11/32 cases) [3], and 23% in 2026 (3/13 cases). Currently, no specific treatment or cure is available for hantavirus infections; clinical management remains supportive, while preliminary trials for targeted therapies have only recently begun [29]. The precise mechanisms of ANDV transmission remain uncertain, with direct contact, droplets, aerosols, and contaminated fomites all proposed as possible routes. Available evidence suggests that transmission requires close and prolonged contact, while long-range aerosol transmission is unlikely [13, 19, 30].

Unlike COVID-19, where older age was associated with increased fatality risk [31], there is currently no evidence that age is a determinant of HPS infection or mortality. During the 1996 outbreak, infected individuals had a mean age and interquartile range (IQR) of 38 [13; 70] years [15]; in 2018, 38 [27; 58] years [13]; and in 2026, based on available data, we obtained 64 [58; 68] years. These differences likely reflect the demographics of the affected populations rather than differences in susceptibility; for instance, the *MV Hondius* mainly hosted middle-aged (45-65 y/o) passengers (The Guardian, “MV Hondius: the ice-breaking expedition cruise hit with hantavirus cases”, 2026/05/05). Also, fatalities were similarly distributed across age groups. In 1996, the six deceased patients with available age data had a mean age of 40 [30; 50] years, while the remaining three were active workers likely within a similar age range [15]. In 2018, deceased patients had a mean age of 45 [20; 70] years [13]. The three deaths in the 2026 outbreak occurred among individuals aged 65, 69, and 70 years. Therefore, current evidence does not support an increased risk of ANDV transmission or fatality associated with age, although further investigation is needed.

### 2.2 Estimation of epidemiological parameters

Reliable outbreak models and plausible scenarios require accurate estimates of epidemiological parameters and of their associated uncertainties. Social gatherings and cruise ship environments are often represented as temporal multilayer small-world networks with community structure [32, 33, 34]. However, the limited availability of transmission trees, contact patterns, and detailed outbreak information for all three ANDV outbreaks prevents the calibration of such complex models. We therefore adopt a simpler but informative approach, which requires little assumptions and is amenable to detailed analysis: a SEIR (Susceptible, Exposed, Infectious, Removed) model embedded in a prototypical small-world network to represent social contacts and go beyond unrealistic well-mixing assumptions [35]. This framework introduces a limited number of long-range interactions while preserving clustering among close contacts, requires few parameters to calibrate, and enables investigation of the interplay between biological and network effects. Small-world models have previously been used to describe real-world social networks and gatherings [36], making them a suitable representation of the heterogeneity and stochasticity of viral spread. The rationale for model selection and its implementation are described in Methods.

Model parameters are related either to network structure or to epidemic dynamics. For the network, we set the default number of nodes to *N* = 150, corresponding to the number of passengers aboard the cruise ship (according to *Oceanwide Expeditions*’ report), with rewiring probability *p_r_* = 0.1 and average degree ⟨*k*⟩ = 6. The latter two values are informed estimates based on standard modelling practices and qualitative observations (see Methods). Sensitivity analyses were performed to assess the reliability of the fitted epidemiological parameters, whose effects on outbreak scenarios are further explored below.

The SEIR model parameters (Fig. 2, black) are the transmission rate *β*, incubation rate *α* (or reciprocal incubation period *T_α_*) and recovery rate *γ* (or reciprocal infectious period *T_γ_*). The average reproduction number is defined as:

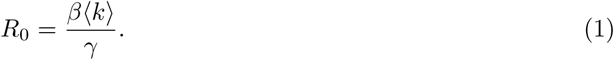

**Figure 2:**
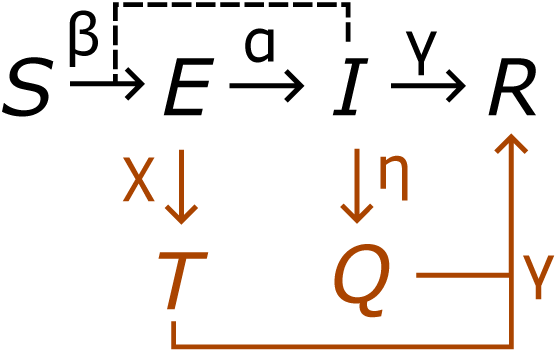
Scheme of the considered states (Latin letters) and of the rates governing the probability of transition from one state to another (Greek letters). In black, the rates and states of the base model; in orange, those of the model augmented with control strategies. *T* stands for Tested, *Q* for Quarantined; see Methods for details.

The limited information available for the 2026 outbreak prevents simultaneous estimation of all parameters, and parameter non-identifiability could reduce the reliability of subsequent scenario analyses. Therefore, assuming that transmission and infection mechanisms have remained largely unchanged across outbreaks, we fix parameters where possible, based on previous studies. The median incubation period *T_α_* is set at 18 days from the literature [37], while the median infectious period is derived from the 2018 outbreak as described in Methods and in Supplementary Sec. S3, yielding 10 days. Table 1 summarizes parameter values and uncertainty ranges. These median values are used to estimate the 2026 outbreak reproduction number *R*_0_ by fitting the SEIR network model to incidence data through an Approximate Bayesian Computation (ABC) approach (see Methods).

**Table 1:**
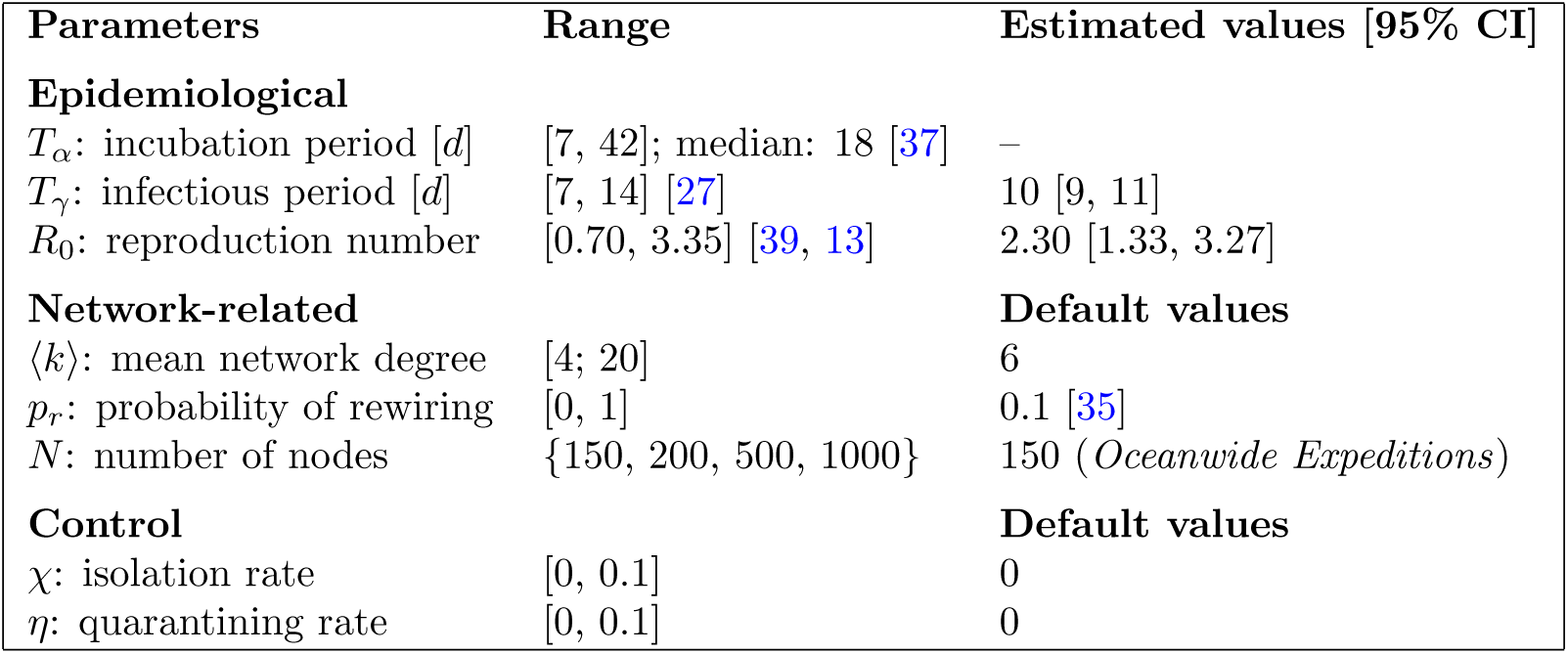
The considered parameters with their values and ranges, used for fitting or for scenario development as explained in the main text. Estimated epidemiological parameters are used as default values when necessary. *T_γ_* and *R*_0_ are estimated independently on two different sets of data. The range and default values for ⟨*k*⟩ is informed by the works of [40, 41, 42, 43] as discussed in Methods. For details about the other parameters, refer to the main text.

We perform the fit over two periods, following the approach of [13] to enable direct comparison with the 2018 outbreak. First, we fit the complete dataset to estimate an average reproduction number for the entire event. We obtain *R*^(*avg*)^_0_ = 1.10 [0.54, 2.58] (median and 95% CI), consistent with the value estimated for 2018 (*R*^(*avg*)^_0_ = 1.19) [13] and with the fit that we also performed on the 1996 outbreak (*R*^(*avg*)^_0_ = 1.60 [0.56, 3.47]). This quantity represents the reproduction number that would have been observed for an outbreak with the same incidence trajectory in the absence of containment measures. Assuming a well-mixed population aboard the ship, it would correspond to a herd immunity threshold of *N_τ_* = *S*_0_(1 − 1*/R*^(*avg*)^_0_) ≃ 14 individuals. We then fit only the data preceding the implementation of mitigation measures in early May (Fig. 1), obtaining the intrinsic reproduction number in a fully susceptible population without interventions. This yields *R*_0_ = 3.28 [0.94, 5.83], which is statistically consistent (*Z* = 0.9) with the pre-intervention estimate for 2018, *R*_0_ = 2.12 [1.24, 3.35] [13].

These results have three main implications. First, they support the hypothesis that ANDV epidemiological characteristics have remained largely unchanged across outbreaks, as reproduction numbers are consistent within uncertainty intervals. Second, the estimated *R*_0_ = 3.28 [0.94, 5.83] indicates a diffusion potential comparable to that of early COVID-19 variants, for which *R*_0_ exceeded 2.5 [38]. The ABC analysis is also consistent with sustained human-to-human transmission and difficult to reconcile with assumptions based solely on repeated zoonotic exposure. Previous outbreaks could potentially have been explained by repeated rodent exposure combined with long incubation periods rather than sustained transmission [20]; however, the 2026 outbreak included cases appearing beyond the maximum reported incubation period of 42 days. Furthermore, the posterior probability of having *R*_0_ less than 1 is only *P* (*R*_0_ *<* 1) = 0.08, suggesting that the observed cases are unlikely to represent a declining chain of infections caused solely by multiple contacts with external exposures. Third, the difference between *R*_0_ and *R*^(*avg*)^ confirms that mitigation measures are necessary to reduce the reproduction number to an eradication level. Without such interventions, the probability of a larger and potentially more severe outbreak would have been substantially higher.

Finally, we perform a synthesis of the 2018 and 2026 estimates (see Methods) to obtain a more precise characteristic reproduction number for ANDV. We estimate *R̂*_0_ = 2.30 and *σ̂* = 0.49, corresponding to a 95% CI in [1.33, 3.27]. The final network and epidemiological parameters are summarized in Table 1 and used for subsequent sensitivity, uncertainty, and scenario analyses.

### 2.3 Uncertainty in ***R*_0_**

For epidemics spreading on small-world networks, no exact herd immunity threshold exists. For moderate rewiring probabilities *p_r_*, as in our model, the threshold is slightly below the well-mixed SEIR value *N_τ_* = *S*_0_(1 − 1*/R*_0_), which provides an estimate of the average number of infections after which an epidemic would spontaneously decline [44, 45]. Moreover, epidemics generally spread more slowly on small-world networks than on fully connected or random networks [36, 46, 47]; together with the presence of immune individuals disrupting susceptible connections, this observation contributes in qualitatively explaining why ANDV outbreaks have remained limited in size (Fig. 1). However, the stochastic nature of network epidemics requires simulations informed by ANDV-specific parameters to test these hypothesis quantitatively, to predict medium- and long-term dynamics and to evaluate sensitivity to parameters potentially affected by viral evolution. We therefore perform Monte Carlo simulations of the model as described in Methods.

We first investigate the impact of uncertainty in *R*_0_. From the previous analysis, we estimated *R*_0_ = 2.30 [1.33, 3.27] (Table 1). This corresponds, on average, to a herd immunity threshold of *N_τ_* ≃ *S*_0_ · 0.57; for the *MV Hondius*, in the absence of mitigation measures, approximately 80 infections would be expected before spontaneous epidemic decline. However, stochastic effects and uncertainty in *R*_0_ can lead to substantially different outcomes. Figure 3a shows Monte Carlo simulations sampled from the estimated *R*_0_ range. In most scenarios, the epidemic develops a clear peak of approximately 13–15 infections, occurring 4 to 6 months after the index case and eventually affecting most of the population (Fig. 3b). Conversely, when *R*_0_ lies near the lower end of the uncertainty interval, outbreaks may fail to establish due to network effects. The main consequence of stochasticity is therefore a bimodal attack-rate distribution, with a substantial fraction of simulations resulting in no sustained outbreak (Fig. 3b, right). This finding may help explain differences among the 1996, 2018, and 2026 outbreaks. While the 2018 and 2026 events were contained through early interventions such as contact tracing and isolation, the 1996 outbreak may have been limited without explicit control measures because its effective *R*_0_ was near the lower end of the plausible range, potentially influenced by the underlying social network structure (recall that ⟨*k*⟩ explicitly tunes *R*_0_, see Eq. 1). The mitigation effects of the network structure are deeply investigated in a later section.

**Figure 3:**
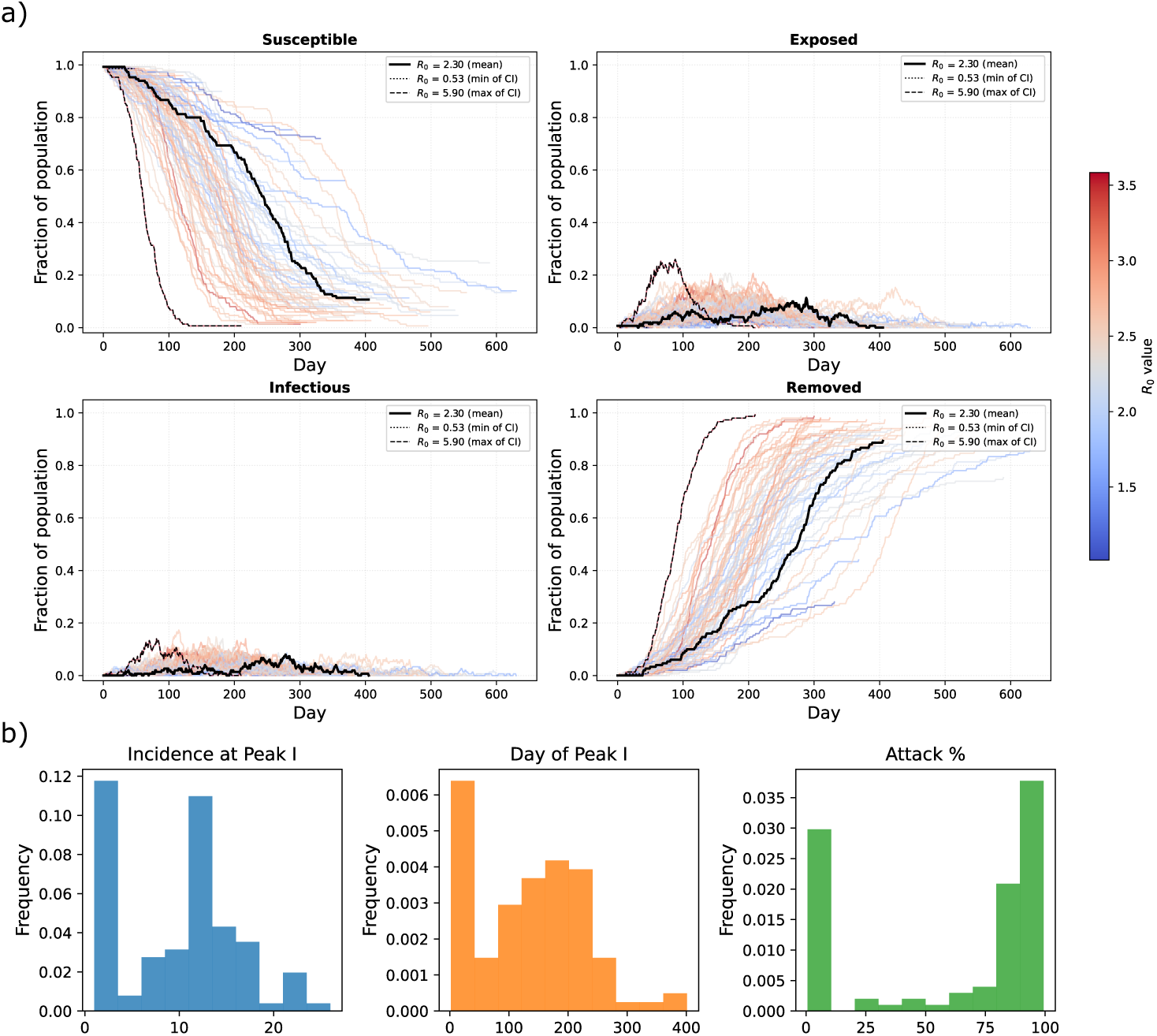
Effects of the uncertainty of *R*0. a) Trajectories of the *S*, *E*, *I* and *R* states, obtained with Monte Carlo simulations of the network model sampled from a Gaussian uncertainty interval in [1.33, 3.27] (*cf.* Methods). Two additional trajectories, associated with extreme *R*0 values described in literature [13, 35], are included for comparison. Note that, for *R*0 = 0.53, the outbreak immediately vanishes. b) Distribution of the ensemble of trajectories: maximum incidence at peak, timing of the peak, and total attack rate.

### 2.4 Uncertainty in the incubation period

The ANDV incubation period *T_α_*is highly uncertain [37], with estimates ranging from 7 to 42 days and a median of 18 days. From Eq. 1, *T_α_* does not determine the initial epidemic growth rate or the herd immunity threshold; however, it affects epidemic timing, including the peak occurrence, overall duration, and required quarantine period. We therefore explicitly account for uncertainty in *T_α_*. Figure 4 shows the resulting trajectories for each epidemic state. As observed for uncertainty in *R*_0_, some outbreaks fail to establish due to network effects, while successful outbreaks display variability in peak size and timing before infecting most of the network. Thus, variation in incubation times contributes to the heterogeneity of epidemic dynamics.

**Figure 4:**
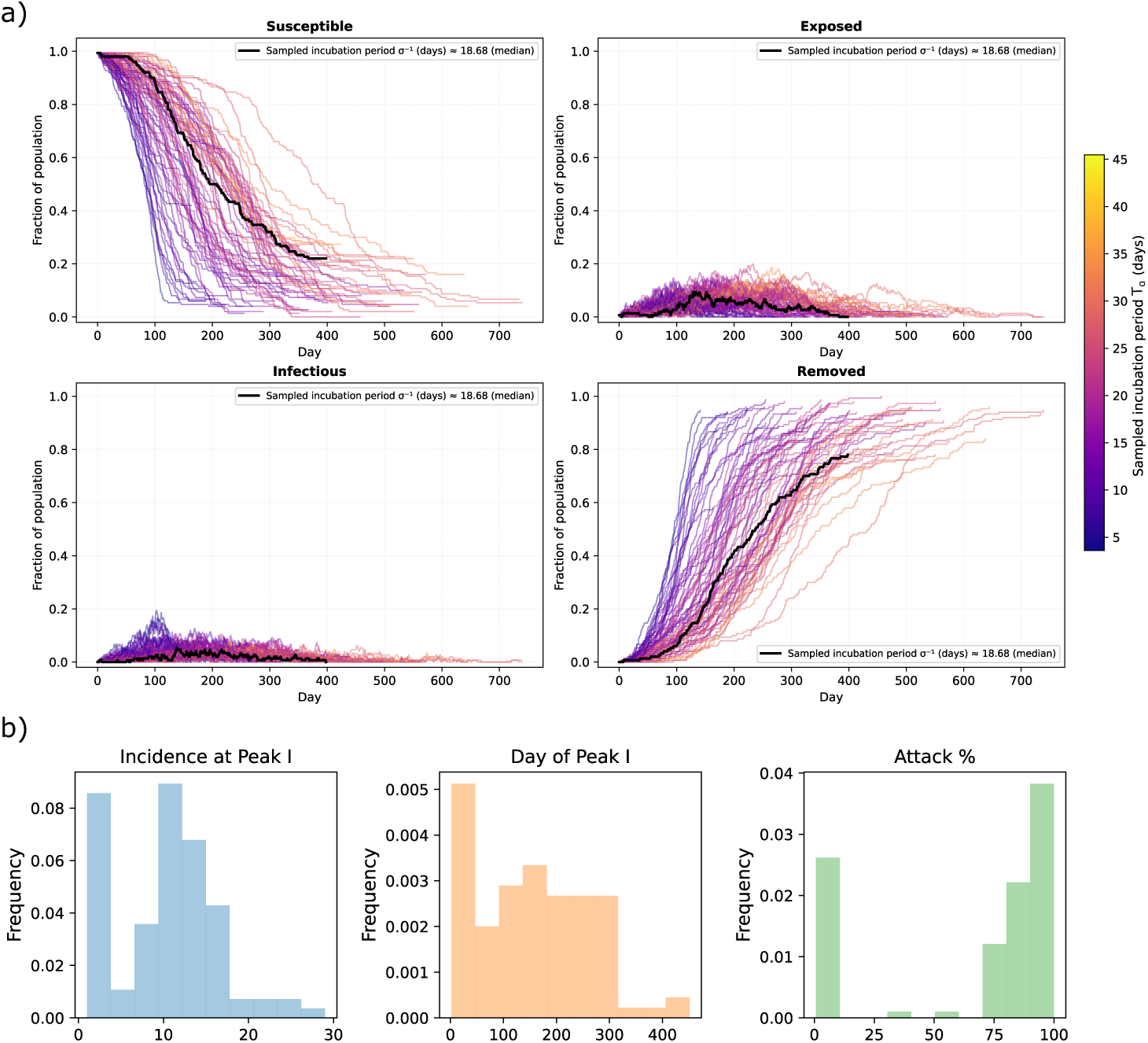
Effects of the uncertainty of *Tα*. a) Trajectories of the *S*, *E*, *I* and *R* states, obtained with Monte Carlo simulations of the network model sampled from a Gamma distribution with 95% CI in [7, 42] (*cf.* Methods). b) Distribution of the ensemble of trajectories: maximum incidence at peak, timing of the peak, and total attack rate.

To quantify whether this heterogeneity differs from that caused by the uncertainty in *R*_0_, we perform Kolmogorov–Smirnov (KS) tests to compare the distributions obtained from the *R*_0_ and *T_α_* uncertainty analyses (Fig. 3b and 4b). For all three considered metrics (peak incidence, peak timing, and attack rate), the pairwise tests yield *p*-values *>* 0.05, indicating no statistically significant differences between the distributions. This suggests that uncertainty in the incubation period is largely captured by uncertainty in *R*_0_. Therefore, when estimating ANDV outbreak progression and peak dynamics, decision makers may rely primarily on accurate characterization of *R*_0_, despite limited knowledge of *T_α_*. However, accounting for the possibility of incubation periods up to 42 days remains essential to define appropriate quarantine durations, as implemented in previous outbreaks.

The uncertainty analysis is complemented by a sensitivity analysis, as described in Methods. While the uncertainty analysis investigates how epidemic trajectories change when infectious individuals have variable and uncertain incubation periods, the sensitivity analysis explores how different epidemics evolve when characterized by different incubation periods and stochasticity arises only from the network structure. Results are reported in Supplementary Sec. S4; both approaches produce comparable epidemic dynamics. Furthermore, the KS test confirms that the distributions of peak size, peak timing and attack rate are not significantly different from those obtained by varying *R*_0_, suggesting that uncertainty and sensitivity on *T_α_*can be effectively incorporated through uncertainty in *R*_0_, thus reducing model complexity, when developing outbreak scenarios.

Finally, we investigate whether incubation-period uncertainty can be represented using an all-to-all compartmental ODE SEIR model with delay, following the approach of [48]. In this framework, the *E* → *I* transition is governed by an Erlang-distributed delay, a special case of the Gamma distribution that connects the model to renewal equation approaches [49]. Details on the delayed SEIR implementation, its connection with previous studies, and results are provided in Supplementary Sec. S4. This analysis confirms that uncertainty in the incubation period primarily affects the timing of the epidemic peak. It also shows that using deterministic approaches produce qualitatively similar, but quantitatively different results than using a network dynamics, providing a bridge between networkbased and deterministic modelling approaches and supporting the development of future models for ANDV transmission and control.

### 2.5 Uncertainty in transmissibility and superspreading events

Martínez and colleagues [13] suggested that ANDV may exhibit superspreading events, in which a small number of individuals infect far more contacts than average. Such events reflect heterogeneity in transmission that cannot be captured by the mean reproduction number alone [50]. Instead, transmission variability is described by an overdispersed, right-skewed distribution of secondary infections, whose variance exceeds its mean [39]. Superspreading has played a major role in the dynamics of COVID-19, SARS, and MERS [51, 52, 53]. Here we isolate the effect of transmission heterogeneity by keeping the contact network fixed while varying the probability of transmission. The complementary effect, of changing the network structure by increasing the number of contacts [54], is examined later. We model transmission heterogeneity by drawing the infection probability from a Gamma distribution (see Methods), parametrised by the overdispersion parameter *k_d_*, which controls the variance around a fixed mean *R*_0_. A sensitivity analysis is performed around *k_d_* = 0.394, estimated from the 2018 outbreak (Supplementary Sec. S1). The explored range spans values comparable to those reported for COVID-19 [55, 56]. Simulations are repeated for the mean estimate of *R*_0_ and for the bounds of its confidence interval (Tab. 1).

Results from repeated Monte Carlo simulations (see Methods) are shown in Fig. 5. As expected, superspreading events are most frequent at low *k_d_*, where transmission is highly heterogeneous, and become progressively rarer as *k_d_* increases (top row). For *k_d_* = 0.04, most transmission events occur via superspreading, while *k_d_* = 5 generates lower heterogeneity and most infectious nodes produce about *R*_0_ new cases. Larger *R*_0_ values consistently generate more superspreading events, indicating that such events are plausible for ANDV, similarly to what was observed in previous outbreaks.

**Figure 5:**
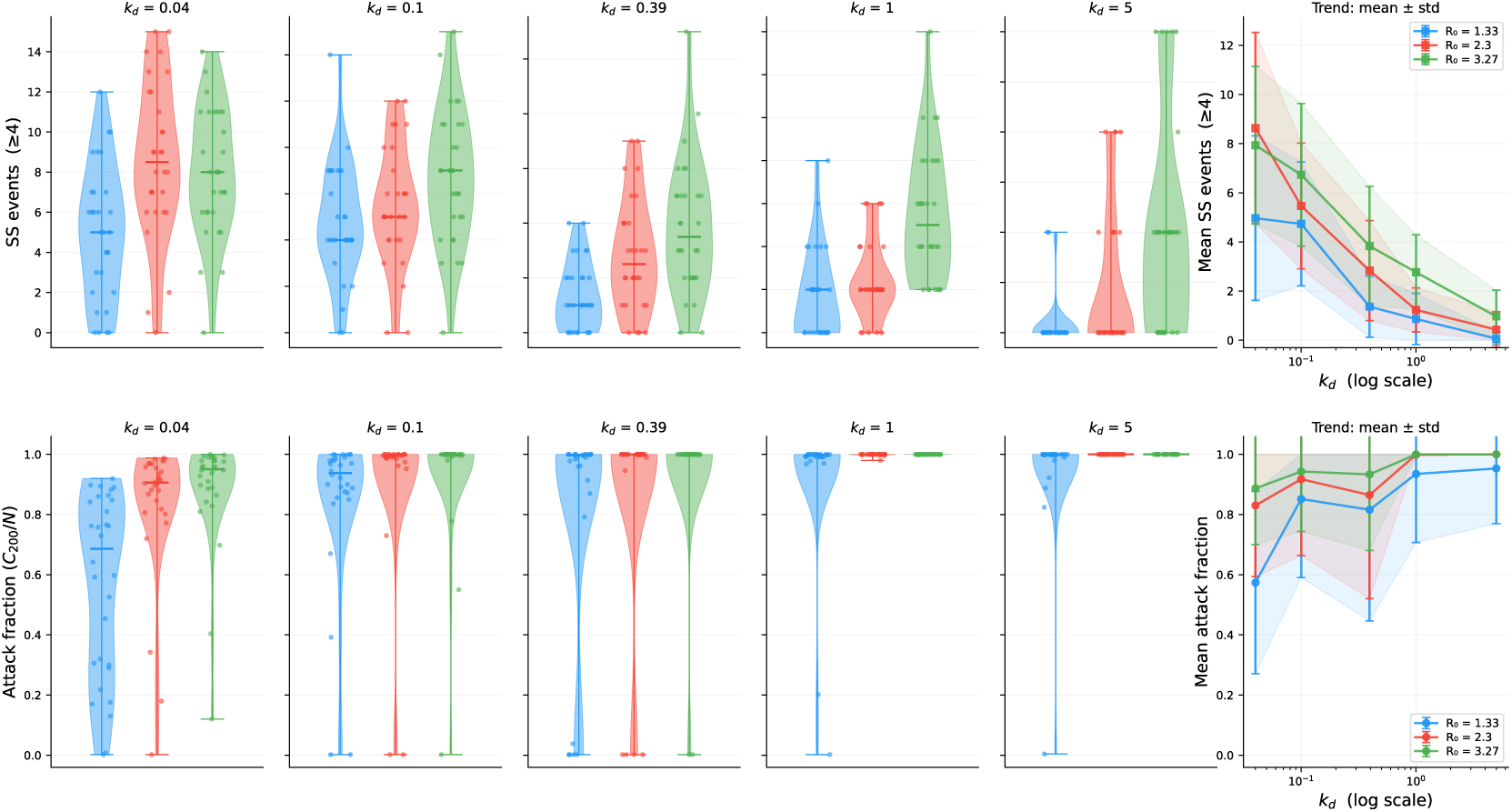
Effect of uncertainty in transmission on epidemic dynamics. Top row: Distributions of superspreading events depending on *k_d_* and *R*_0_ values. Bottom row: The simulated attack fraction (cumulative count of cases over total population) after 200 days, depending on *k_d_* and *R*_0_ values. In both rows, the last panel summarises the trends of mean and one standard deviation.

To assess the severity of epidemic progression, we measure the attack rate after 200 days (approximately 5 months), taken after the median peak time observed in Figs. 3b and 4b. The 200-days attack rate (bottom row) remains high across all *R*_0_ values, except for occasional runs at low *k_d_*. Strong overdispersion increases not only the probability of superspreading but also the probability that infected individuals generate few or no secondary cases. These opposing effects balance each other to preserve the same mean *R*_0_, while increasing the chance of stochastic extinction. Outbreaks with greater transmission heterogeneity are thus slightly more likely to die out early, reducing the average attack rate. Superspreading therefore complicates outbreak management and contact tracing but does not necessarily lead to larger epidemics; under some conditions, it can even produce slightly smaller outbreaks. This fact can help explain why rural ANDV spillovers frequently burn out naturally. These early extinctions become more frequent at lower *R*_0_.

### 2.6 Mitigation scenarios following control strategies

Mitigation and suppression of epidemic outbreaks can be achieved through contact tracing, isolation of exposed individuals, or quarantine of infectious cases [57, 58]. The effectiveness of these measures depends on the basic reproduction number, the implementation delay, or the intervention strategy. We now assess simple control measures for ANDV transmission on a small-world network using the estimated *R*_0_. Because real-world testing and tracing capacities are difficult to quantify [25], we assume that exposed individuals are identified (via testing or contact tracing) and isolated at rate *χ*, and infectious individuals are quarantined at rate *η* (see Fig. 2, in orange, and Methods). Consistent with the 2018 and 2026 events, interventions are assumed to begin *t*_int_ = 30 days after the first exposure (see Methods). If the dynamics occurs on a homogeneous population, the effective reproduction number of a SEIR model after interventions becomes [57, 58]:

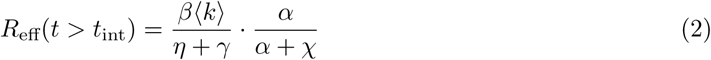

Setting *R*_eff_ *<* 1 yields the intervention thresholds, above which an epidemic can be surely suppressed. If contact tracing is particularly challenging and is therefore absent, *χ* = 0 and *η_c_* = *γ*(*R*_0_ − 1). For the median ANDV parameters, this gives *η_c_* = 0.13, corresponding to quarantine within about 7 days. For comparison, *η_c_* in case of the COVID-19 epidemic (using the median parameters for the alpha variant [58, Table 2]) was 0.51, corresponding to quarantine within about 2 days: a much more stringent requirement for containment. If, instead, quarantine is not feasible (*η* = 0) and tracing and isolation are preferred, *χ_c_* = *α*(*R*_0_ − 1). In this case, knowing the incubation period is crucial to establish a rollout strategy of tracing and isolation. For the median value *α* = 1*/*18 days*^−^*^1^, *χ_c_* = 0.07: to curb an infection, isolation should occur within about two weeks. For the alpha variant of SARS-CoV-2, *χ_c_*was 0.3: isolation was required to occur within 3 days. We therefore see that the current ANDV variant is much more controllable than SARS-CoV-2 even on a homogeneous population.

To fully account for network heterogeneity, we again employ Monte Carlo simulations. The simulations are performed (30 replicates for each combination of *χ* and *η*) on networks of 500 nodes to enable comparison with the subsequent analysis on network effects. Figure 6a shows the effect of combining *χ* and *η* to reduce *R*_eff_ from Eq. 2: the light grey line marks the values for which *R*_eff_ = 1. Instead, Fig. 6b shows the effect of control measures in suppressing the curves. As above, we measure the attack rate at 200 days as reference. Figure 6 also uses the same colormap range as Fig. 8. Owing to stochasticity and network effects, epidemic suppression can occur even below the theoretical thresholds. As a reference to interpret the rate values, during the first wave of COVID-19, Denmark activated an equivalent quarantine rate of *η* = 0.014 on top of social distancing mandates, while Austria had a *χ* = 0.001 trace-and-isolate rate [58]. Finally, we observe that test-and-isolate rates are more effective than quarantine rates with the same values in curbing an ANDV epidemic: while quarantine of infectious individuals is a necessary step, contact tracing should also be performed actively.

**Figure 6:**
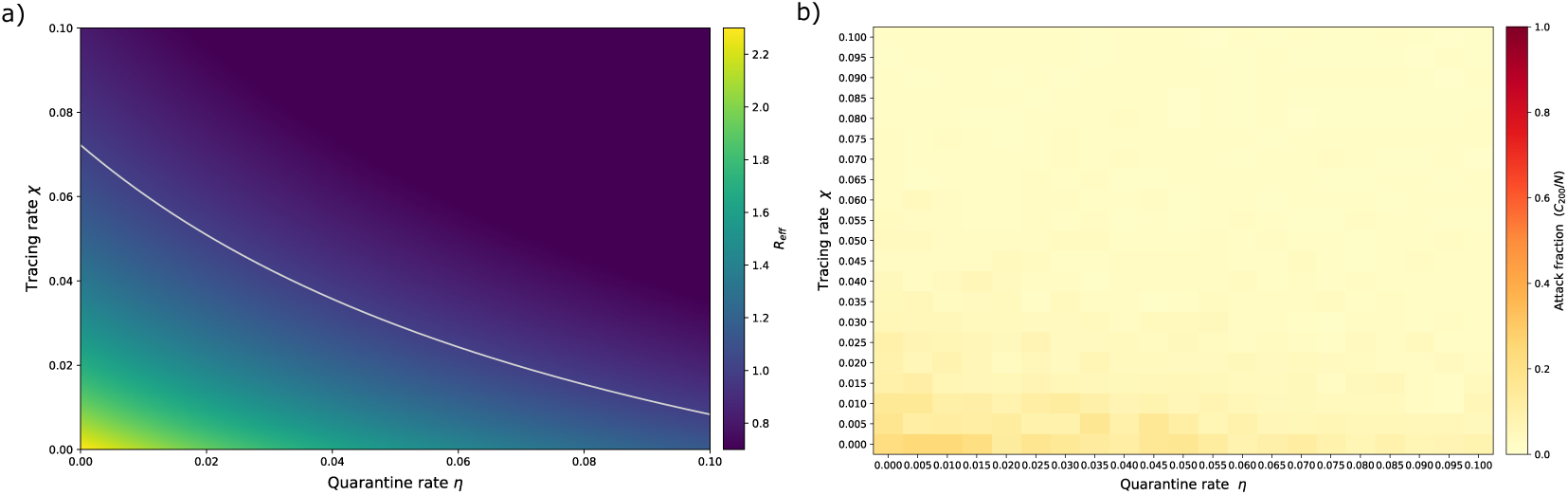
Effect of testing-and-isolation rate *χ* and quarantining rate *η* on epidemic dynamics. (a) Theoretical effect on *R*_eff_, following Eq. 2. The light grey line identifies critical values for which *R*_eff_ = 1. (b) The simulated attack fraction (cumulative count of cases over total population) after 200 days.

### 2.7 Asymptomatic infectious carriers may hinder the efficacy of quarantine

There is currently no evidence that asymptomatic individuals transmit ANDV; horizontal transmission has only been linked to symptomatic cases. However, viral RNA is detectable in blood 5–15 days before symptom onset, suggesting that pre-symptomatic infectiousness cannot be completely excluded [19]. If asymptomatic individuals were infectious, epidemic dynamics and mitigation strategies could change substantially. This scenario represents either the emergence of a variant capable of pre-symptomatic transmission or an unrecognised feature already observed in other hantaviruses [21].

To investigate this, we extend the network SEIR model with quarantine by adding an Asymptomatic (*A*) compartment. A fraction *θ* of exposed individuals becomes symptomatic infectious (*I*), while (1−*θ*) enters *A*. Pre-symptomatic infection is modelled as transmission between nodes in *A* and *S*, occurring with the same *β* as in case of symptomatic infection. Individuals remain in *A* for a period *T_ζ_* before developing symptoms and moving to *I*. The SEIARQ model therefore introduces two parameters: *θ* relates to asymptomatic seroprevalence, which varies greatly between regions, hantavirus species and exposure to rodents, from 1% in US [59] to 40% and beyond in South American indigenous populations [60, 61]; and *T_ζ_*, the duration of pre-symptomatic infectiousness. We assume that quarantine acts only on symptomatic cases and we evaluate how *η*, *θ*, and *T_ζ_* influence epidemic spread and control.

Like before, Monte Carlo simulations are averaged over repeated runs. We first vary *θ* between 0 (all asymptomatic) and 1 (all symptomatic) holding *T_ζ_* = 10 days, reflecting the average interval of viral RNA detection before symptoms [19]. Separately, *T_ζ_* ranges from 1 to 18 days (the median incubation period) with *θ* = 0.5 (half-and-half split). All other parameters are kept at default values. Figure 7 reports the attack rate after 200 days, depending on the asymptomatic parameters and the quarantine rate. In Fig. 7a, *θ* determines what fraction of infections are symptomatic and are therefore visible and quarantinable. When *θ* is small, most infections route through the silent *A* state, and even large *η* cannot prevent a large epidemic. Quarantine becomes effective only when more than one third of the infectious population is symptomatic. In Fig. 7b, increasing *T_ζ_* prolongs silent transmission before detection, reducing quarantine effectiveness. However, its impact is weaker than that of *θ*, indicating that the proportion of transmissible asymptomatic infections is the dominant uncertainty and should receive greater attention in preparedness planning.

**Figure 7:**
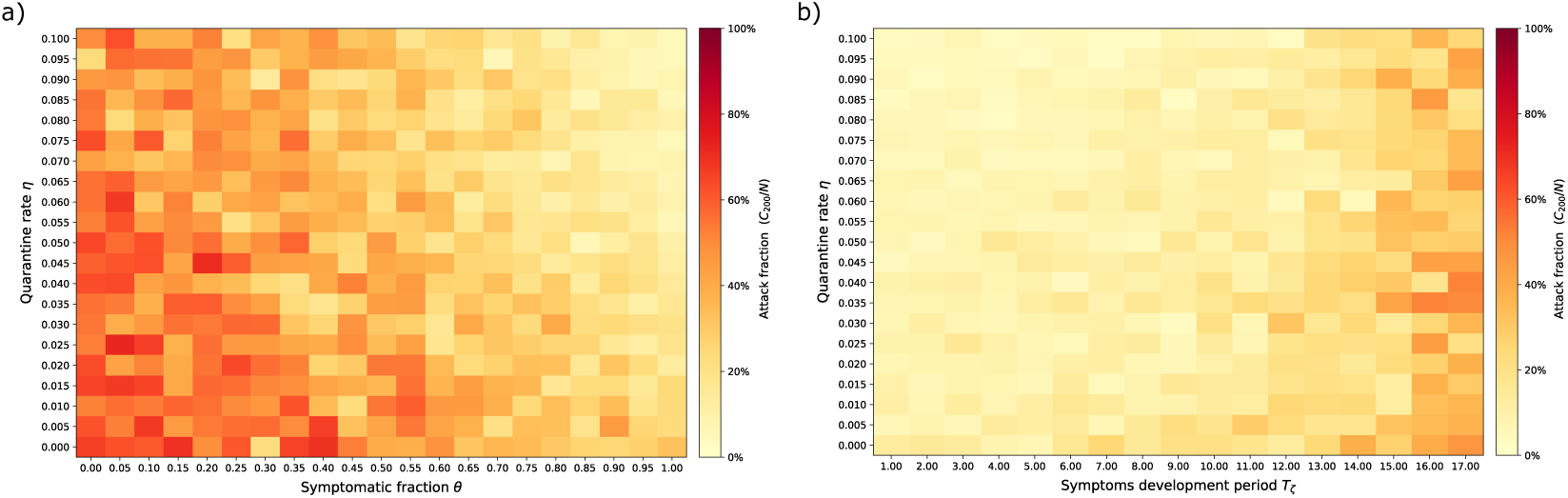
Effect of quarantining rate *η* on the attack fraction at 200 days, depending on symptomatic fraction and symptoms development period. (a) Dependence on *η* and *θ*. (b) Dependence on *η* and *T_ζ_*.

Overall, this analysis recalls that quarantine is only as effective as case detection. Undetected asymptomatic infections can sustain transmission and reduce intervention efficacy, while longer *T_ζ_* extends the silent spreading phase. *T_ζ_* is “long” relative to *T_α_*: how much time healthcare professionals have to intervene and trace pre-symptomatics depends on the incubation period and its uncertainty. Although pre-symptomatic transmission does not change *R*_0_ if infectiousness matches that of symptomatic cases, it can substantially weaken quarantine-based control strategies.

### 2.8 Network structure determines epidemic progression

Previous scenarios investigated the effects of epidemiological parameters and control strategies based on testing and isolating infected individuals. We now examine the role of network structure, focusing on the influence of the average degree ⟨*k*⟩ and rewiring probability *p_r_* on epidemic dynamics. We again measure the attack fraction after 200 days. We perform Monte Carlo simulations (30 replicates for each combination of ⟨*k*⟩ and *p_r_*) on networks of 500 nodes to explore a wider range of connectivity values. Epidemiological parameters are fixed at their median estimates (Table 1). Figure 8 summarizes the mean attack rate at 200 days, while example trajectories are reported in Supplementary Sec. S5. For very low ⟨*k*⟩ or *p_r_*, attack rates remain limited because the network is highly sparse, with individuals interacting mainly within small and close groups. Long-range transmission events become rare, and outbreaks fail to propagate beyond local clusters because infectious individuals quickly exhaust their available contacts. Reducing long-range interactions can therefore substantially slow transmission, as these rare shortcuts are often responsible for cross-cluster spread. This scenario represents an ideal scenario where spontaneous or informed social distancing is effective: here, limited mixing and absence of long-range contacts suppress outbreaks without additional interventions. Such a situation may help explain the small ANDV clusters observed in the early 2000s [2], which remained confined within families or close social groups due to limited external contacts.

Increasing *p_r_* increases population mixing and approaches a random network as *p_r_* approaches 1, by introducing more connections among people that are not spacially or socially proximal. Similarly, increasing ⟨*k*⟩ gives each individual more potential transmission pathways through more neighbours. Increasing both parameters moves the system towards a well-mixed regime, typically represented by compartmental SEIR models, where individuals interact with many others as in large gatherings or highly mixing environments, or where airborne viruses like SARS-CoV-2 diffuse in confined spaces. Under these conditions, attack rates increase, although not monotonically due to network stochasticity, eventually producing large and rapid outbreaks with attack fractions close to one.

Thus, even with identical epidemiological parameters, the network structure strongly influences epidemic outcomes. This influence would not have been captured by branching process models [25] that cannot directly evaluate the impact of interventions targeting social connectivity. Instead, out network model highlights that social structures that limit contacts, preserve local clusters, and reduce long-range mixing can delay or suppress outbreaks, reduce peak burden on healthcare systems, and provide additional time for intervention. Such effects are particularly relevant for ANDV, for which long-distance transmission appears limited. Rapidly identifying and isolating local clusters around index patients is thus an effective strategy for ANDV containment. Figure 8 also provides information for targeted and custom interventions, depending on the conditions where an outbreak first develops. For instance, contacts on cruise ships are, on average, above 12, especially in dining halls or sport facilities [40]; an immediate intervention against ANDV diffusion would require reduced access to such places (to reduce ⟨*k*⟩ directly) or the mandate to only engage in social activities with close friends or families, to reduce *p_r_* and the chance of spread to external clusters. For gathering events, where 5 to 7 close contacts are usually observed [41], attention should be dedicated to avoid long-range contacts and mixing among groups, rather than banning the events altogether.

Finally, the dashed blue line in Fig. 8 indicates *p_r_* = 0.1, the default value used in our simulations because it is representative of small-world social networks [35]. For this value, epidemic outcomes show limited sensitivity to variations in ⟨*k*⟩, further supporting the robustness of the fitted network parameters.

**Figure 8:**
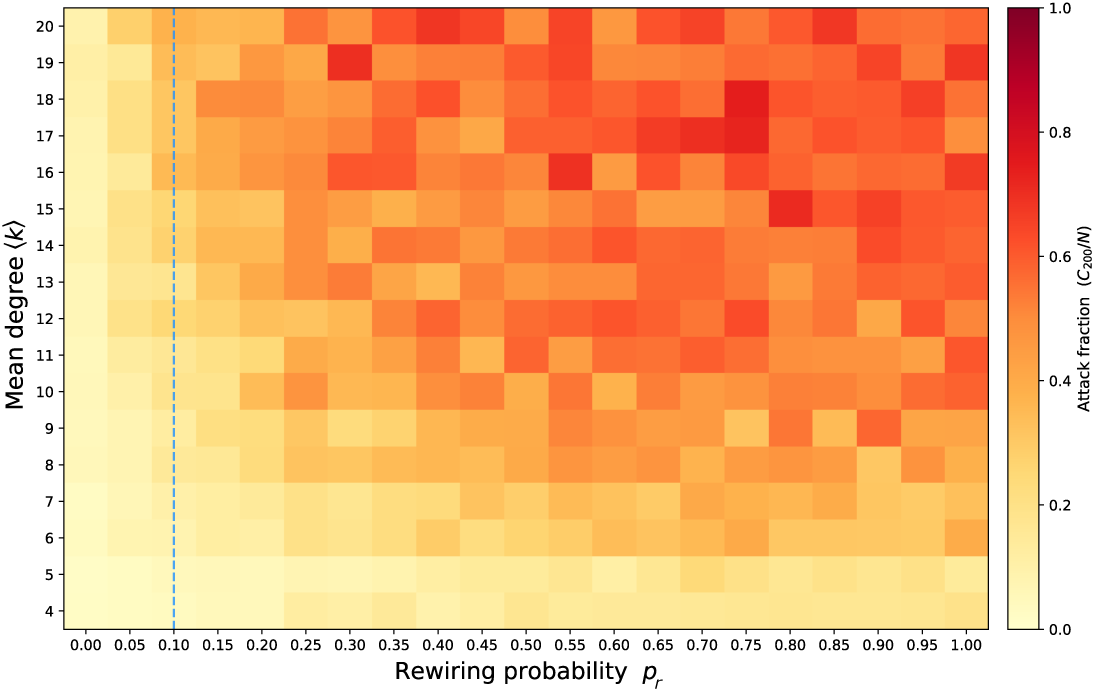
The attack fraction (cumulative count of cases over total population) after 200 days, depending on networks with increasing ⟨*k*⟩ and *pr* . The blue line corresponds to *pr* = 0.1, used for the fit.

## 3 Discussion

This work combines evidence from historical *Andes* virus (ANDV) outbreaks with data from the recent *MV Hondius* event to assess the risk of human-to-human transmission and identify conditions favouring future outbreaks, with the aim of developing preventive scenarios. Fitting a representative network model confirms sustained human transmission while indicating that the virus has not substantially changed its outbreak potential over the past 30 years. The estimated overall reproduction number is *R*_0_ = 2.30 [1.33, 3.27], comparable to the original COVID-19 strain [58], implying that ANDV is unlikely to disappear without interventions such as distancing, isolation, or quarantine. This estimate provides a reference value for future hantavirus modelling.

Using Eq. 1, the estimated *R*_0_ corresponds to an average transmission probability *β* between 1% and 6% for realistic contact networks in social events or ships, since the range of average contacts reasonably varies between 4 and 20 (see the range of ⟨*k*⟩ in Tab. 1), or between 0.6% and 8% across the full uncertainty range of *R*_0_. These values agree with empirical estimates, ranging from below 3% in households to about 18% among sexual partners [19], and are comparable to reported secondary transmission probabilities for COVID-19, which ranged between 0.1% on public Chinese transportation and 10.3% on households [62]. The uncertainty in transmission and the chance of superspreading are also comparable to those estimated for COVID-19 outbreaks worldwide: *k_d_* = 0.394 is close to *k*^(COVID)^ = 0.41[0.23; 0.60] [50], having extremes at 0.04 [55] and around 3 [56], meaning that 2 to 20% of infectious cases likely caused more than 80% of transmission.

Therefore, compared to COVID-19, ANDV may exhibit transmission potential within the same order of magnitude, captured by similar *R*_0_, *β* and *k_d_* values. ANDV preparedness would thus greatly benefit from lessons learned during the COVID-19 pandemic. The main difference lies in the transmission mode. COVID-19 spreads through aerosols and is strongly influenced by ventilation, occupancy, masking, and exposure time [63]; this is usually well captured by models of dense networks with a high number of neighbours. Instead, current evidence indicates that ANDV requires close contact. Consequently, contact-network structure is more important than small variations in epidemiological parameters. Surveillance should therefore prioritise mutations enabling aerosol or long-range transmission, which could greatly increase outbreak potential.

The current transmission characteristics also make ANDV more amenable to control. The sparse transmission structure makes it potentially easier to isolate clusters of proximal individuals and break the chain of infections; the network-related stochasticity helps suppress the epidemic at values of *R*_0_ higher than one; due to long incubation and infection periods, isolation of exposed individuals or quarantine of infectious cases can be effective even if slower than what was required to suppress COVID-19. These findings help explain why past outbreaks remained localised and support the conclusion of [25] that future outbreaks, having the same conditions as past ones, are unlikely to sustain transmission if timely interventions are implemented.

Our analysis identifies complementary mitigation strategies. Where efficient tracing systems exist, rapid quarantine can effectively suppress outbreaks. However, tracing capacity varies widely across countries: according to the Global Health Security (GHS) Index (https://ghsindex.org/), only a few countries achieve high Early Detection and Reporting (index 4) scores, while the global average remains below 40%. Less prepared countries should therefore aim for social distancing and isolation of clusters via network-based interventions, using our results as guidelines.

The possibility of more diffusive or infectious variants also calls for greater precautions in addressing both epidemiological and network determinants of disease spread. We thus explored scenarios involving uncertainty in epidemiological parameters and viral evolution. Variations in *R*_0_ and incubation time differently affect epidemic timing and attack rate, guiding the selection of operational intervention windows according to epidemic magnitude. In particular, uncertainties in *T_α_*do not alter the risk of viral spread but are relevant for evaluating the effectiveness and timing of control strategies. Works focusing on predicting the virus attack rate can therefore focus on uncertainties on *R*_0_ without adding complexity associated with *T_α_*, while works focusing on the mitigation and suppression of the infectious curve should also consider the uncertainty in *T_α_* and its impact in the containment strategies. We also quantified the impact of transmission uncertainty, including superspreading events. The shape parameter *k_d_* of the transmission distribution controls transmission variance and overdispersion: a higher likelihood of superspreading is offset by a lower probability of moderate transmission. In network models representative of ANDV, this produces occasional superspreading events, consistent with [13], but may reduce the overall attack rate because fewer infections occur through low-contact transmission. Although superspreading remains difficult to manage clinically, it may have the counterintuitive effect of slightly reducing the overall epidemic size. Finally, we have assessed that pre-symptomatic transmission substantially reduces the effectiveness of quarantine, especially when asymptomatic infections outnumber symptomatic ones, making contact-reduction measures essential.

We acknowledge that our study has several limitations. Calibrating the model to historical outbreaks assumes that ANDV transmission has remained stable over time; although supported by statistical analyses, this assumption should be verified through genetic studies of the virus and its potential variants. Incidence dates from the 2026 outbreak are ambiguously reported as symptom onset or as positivity to tests. Since prior studies suggest that symptom onset corresponds to the beginning of infectiousness, and positivity occurs within the uncertainty interval of the incubation period, we thus used the reported dates in the fit and performed an uncertainty and sensitivity analysis; however, consistency in reporting is recommended in future surveillance protocols to reduce uncertainties. Finally, interventions are modelled with constant isolation and quarantine rates, providing a simple and conservative framework [57, 58]. More realistic adaptive or intermittent control strategies [64] may achieve greater mitigation and warrant future investigation.

## 4 Methods

### 4.1 Data acquisition and management

Among the limited but significant HPS outbreaks recorded since the first evidence of human-to-human transmission, three have received substantial scientific and public attention: the 1996 outbreak in southern Argentina, the 2018 outbreak in Epuýen, and the 2026 *MV Hondius* outbreak.

For the 1996 outbreak, we retrieved infection timing, case numbers, and the partially reconstructed transmission tree from [15]. Reported timings corresponded to symptom onset. Information about control measures was unavailable; however, several patients were hospitalized, and late cases were likely associated with hospital exposure, suggesting that some degree of contact reduction occurred.

For the 2018 Epuýen outbreak, case progression and the reconstructed transmission tree were obtained from [13]. Detection times corresponded to fever onset for patients later confirmed by RT-qPCR, as fever was the only prodromal symptom consistently observed. Exposure dates and incubation periods were retrospectively estimated through surveys administered to patients. Control measures were introduced by the Ministry of Health on 31 December 2018. All epidemiological information was obtained from the original study [13].

Between April and May 2026, an HPS outbreak occurred aboard the commercial cruise ship *MV Hondius*. Twelve cases were confirmed through PCR and/or serology, with one additional probable case, who showed signs and symptoms and was epidemiologically linked to a confirmed infection; three individuals died [27]. Infection timing and case information were collected from official public health reports [28, 27] and media news articles: “Hantavirus strain that spreads between humans found in cruise ship passenger” (BBC, 2026/05/06); “Hantavirus cases linked to cruise ship rise to 12 after crew member tests positive” (NBCNews, 2026/05/22); “Spanish national evacuated from hantavirus cruise ship tests positive” (Reuters, 2026/05/25); “3 more passengers, Spanish, French and American, evacuated from cruise ship test positive for hantavirus” (CBC, 2026/05/12); “Canadian cruise passenger isolating in B.C. tests ’presumptive positive’ for hantavirus” (CBC, 2026/05/16); “Man with hantavirus is being treated at Zurich University Hospital” (SwissInfo, 2026/05/06). Timing refers to symptom onset for the confirmed cases. Age information was available for 9 of 13 infected individuals and for all deceased patients. Ship occupancy data were obtained from the official press release of the ship operator *Oceanwide Expeditions*, https://oceanwide-expeditions.com/press/press-update-m-v-hondius-8-may-2026-19-00-hrs-cet. Information on onboard mitigation measures was obtained from interviews reported by the news article “Hantavirus-hit cruise ship heads to spain after three people evacuated” (Reuters): from early May, passengers were instructed to reduce close contacts, wear masks, and frequently use hand sanitizers. Symptomatic individuals were evacuated in Cabo Verde on May 6th, while full disembarkation and repatriation began on May 10th from Tenerife, Canary Islands. No information about passenger-level exposure patterns or about the transmission chain is available.

All epidemiological data were collected and standardized as incidence and timing arrays. No pre-processing was applied before model fitting and analysis.

### 4.2 The base small-world network SEIR model and its parameters

Model development requires balancing realism with the limited availability of epidemiological data. Based on the evidence reviewed in Sec. 2.1, we introduced a small number of assumptions, which are either tested against empirical observations or explored through sensitivity analyses. First, we accounted for disease transmission in small, heterogeneous communities. Available evidence indicates that ANDV spreads through close contacts and does not exhibit substantial airborne diffusion, even in crowded environments such as social gatherings or cruise ships. Therefore, rather than using classical compartmental models based on homogeneous mixing assumptions, which would inadequately represent transmission patterns, we adopted a network-based modelling framework [36, 46]. Supplementary Sec. S4 instead investigates the possibility of using a delay SEIR model to get complementary qualitative insights.

Detailed network or agent-based models have previously been used to reconstruct outbreaks when high-resolution contact data were available, such as COVID-19 transmission on ships or campuses [32, 33], or in simulated epidemic scenarios [34]. However, no detailed contact information is available for the *MV Hondius* passengers or for previous HPS outbreaks. Using such models would therefore increase the number of unidentifiable parameters and introduce additional uncertainty without improving empirical grounding. We therefore adopted the Watts-Strogatz small-world model [35] as a parsimonious but realistic representation of social contacts, given the available data. It represents a compromise between realism and identifiability and captures three key characteristics of human social interactions: local clustering among close contacts, occasional long-range interactions between otherwise unrelated individuals, and path lengths substantially shorter than those of regular lattices. These characteristics are relevant for ANDV transmission and relate to the coexistence of small household clusters and occasional larger outbreaks, such as those observed in 1996, 2018 and 2026.

For parameter fitting and scenario development, we considered a fixed realization of a Watts-Strogatz network G(*N, k, p_r_*), where *N* is the number of nodes, *k* the average degree, and *p_r_*the rewiring probability. Following the standard construction procedure [46], *N* nodes are arranged on a *k*-regular ring lattice, where each node is connected to its *k/*2 nearest neighbours on either side. Each edge (*i, j*) is then independently rewired with probability *p_r_* by connecting *j* to a uniformly selected node *j^′^* ∈*/ i* ∪ N *^′^*(*i*), preserving the mean degree ⟨*k*⟩ = *k*. See Supplementary Sec. S2 for an example.

The choice of ⟨*k*⟩ requires an informed estimate. Studies of cruise ship contacts report approximately 20 unique contacts per day (IQR 10–36), with around 60% (that is, about 12 [6; 21]) occurring in settings such as dining areas or recreational facilities, where prolonged exposure is more likely [40]. Close and prolonged interactions generally involve 2 to 4 individuals [40]. Studies of social gatherings report between 2 and 14 contacts per participant, with approximately 5–7 close contacts [41]. Another survey estimate around 8 average daily contacts in generic settings, which decreased substantially during pandemic restrictions [42]. Wong et al. [43] also suggest that different countries experienced varying numbers of daily contacts, seldom more than twelve and often below 2 if restrictions are in place. Based on these observations, we set ⟨*k*⟩ = 6 as an average and reasonable default value and later evaluate the sensitivity to this assumption.

In the model, each node *i* ∈ N = 1*, . . ., N* is assigned one epidemiological state for each time *t*: *x_i_*(*t*) ∈ {*S, E, I, R*}. These correspond to the SEIR states on a network [34]: susceptible (*S*), exposed and incubating but not infectious (*E*), infectious (*I*), and removed (*R*), including recovered, immune or deceased individuals. The exposed compartment accounts for the long incubation period of hantavirus infections, during which individuals are assumed not to be contagious [37]. The aggregate compartments size at each time *t* is *S*(*t*) = Σ*^N^*_*i*=1_ **1** [*x_i_*(*t*) = *S*] for the Susceptible pool, and analogously for all the others. The total population is conserved at each time step, such that *S* + *E* + *I* + *R* = *N* . The population size *N* is chosen according to the application. For fitting the 2026 outbreak and related scenarios, we use *N* = 150, corresponding to the number of passengers aboard the *MV Hondius*. For the 1996 and 2018 outbreaks, which occurred in small communities, we use *N* = 200 as an estimate of the socially connected population: both events occurred in small villages and not all citizens and visitors had contacts with one another. In both cases, results are largely insensitive to this choice because outbreaks ended before susceptible depletion occurred: repeating the fitting procedure with *N* = 300 and *N* = 500 changed estimates by less than the third significant digit. Scenario analyses were additionally tested with *N* = 500 and *N* = 1000, producing longer and smoother outbreaks but without significant changes in the outbreak statistics.

The system evolves in discrete daily steps, from time *t* to time *t* + 1. In all cases, we begin from initial conditions *S*(0) = *N* − 1, *E*(0) = 1; the remaining states are initially set to 0; the choice corresponds to having a single infectious person (e.g., due to exposure to rodents’ shedding) in a group of susceptible individuals. The simulations stop when no infectious nodes remain. All transitions for node *i* at time *t* depend only on the state of the node, *x_i_*(*t*), and on the states of its neighbours {*x_j_*(*t*) : *j* ∈ N(*i*)}, making the dynamics a Markovian update on the network. The update for the susceptible nodes *x_i_*(*t*) = *S*, to transition into a state *x_i_*(*t*) = *E*, is governed by independent Bernoulli trials per infectious neighbour, according to

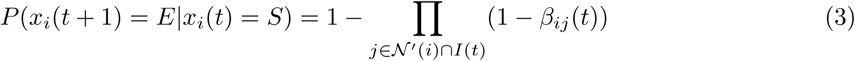

where *I*(*t*) = {*j* : *x_j_*(*t*) = *I*} is the set of currently infectious nodes, and *β_ij_*(*t*) = *β* is the transmission probability from an infectious neighbour *j*. Transition from *x_i_*(*t*) = *E* to *x_i_*(*t*) = *I* follow the standard rule *P* (*x_i_*(*t*+1) = *I*|*x_i_*(*t*) = *E*) = *α*, where *α* [days*^−^*^1^] is the inverse of the incubation period, *α* = 1*/T_α_*; *T_α_* has been estimated to range between 7 and 42 days, with a median of 18 days [37]. Similarly, the transition from Infectious to Removed nodes is governed by *P* (*x_i_*(*t* + 1) = *R*|*x_i_*(*t*) = *I*) = *γ*, where *γ* [days*^−^*^1^] is the inverse of the infectious period *T_γ_*, which was estimated to lie between 7 and 14 days [27]; a more precise median value is fitted, as described below, using data from the 2018 outbreak. *R* is an absorbing state, as there is currently no evidence of waning immunity. As initial condition, we consider all nodes being in the Susceptible state, except for a single exposed node that is randomly drawn: *x_i_*_0_ (0) = *E* for one uniformly random *i*_0_ ∈ N, while *x_i_*(0) = *S* ∀*i* ≠ *i*_0_. This corresponds to a single case infected, e.g., by contact with rodents’ shedding, while other people are susceptible.

### 4.3 ABC fitting procedure

The network SEIR model is fitted to empirical data from the three considered outbreaks by fixing all parameters obtained from literature at their median values, setting a reasonable value for ⟨*k*⟩ as described above, and estimating *R*_0_ through the implicit estimation of *β*. This approach avoids identifiability issues by reducing the estimation problem to a single parameter and prevents overfitting, as the number of available incidence observations remains limited.

To estimate *R*_0_, a simple nonlinear fit with grid search is often used [58].; however, this approach provides only point estimates or uncertainty estimates under restrictive Gaussian assumptions. For complex stochastic epidemics with limited incidence data, Bayesian approaches are more appropriate because they provide posterior distributions and credible intervals for epidemiological parameters [65, 66]. Given the stochastic network structure of our model and the available epidemiological information, we use an Approximate Bayesian Computation (ABC) approach [67, 68] to estimate the posterior distribution of *R*_0_. Given the observed incidence time series **I**_obs_ = (*I*_0_*, I*_1_*, . . . , I_T_*), we aim to infer *p*(*R*_0_|**I**_obs_). The likelihood *p*(**I**_obs_|*R*_0_) cannot be computed analytically because the forward model is a stochastic process evolving on a network; therefore, standard Bayesian inference is not applicable. ABC overcomes this limitation by approximating the posterior through repeated simulations without requiring an explicit likelihood function. To reduce dimensionality and avoid non-identifiability, all parameters except *R*_0_ are fixed to their literature-based median values. The observed incidence time series is used directly as the input time series **I**_obs_, and simulated trajectories are compared with observations through the normalized root mean square error (RMSE):

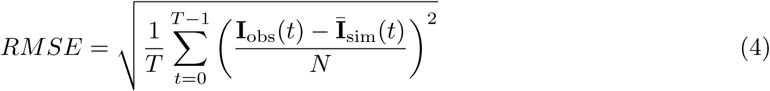

The normalization by *N* makes the metric scale-free in [0, 1], regardless of population size, and comparable across datasets. As a prior, we use a flat and uninformative prior over an epidemiologically plausible range, informed by previous calculation of *R*_0_ from the literature: *R*_0_ ∼ *U* (*R*_min_*, R*_max_), with *R*_min_ = 0.5 and *R*_max_ = 6.0; to allow for some variability associated with the current outbreak, we use extreme values for *R*_0_ that are lower and higher, respectively, than the range from 1 to 5 recorded in previous studies [13, 15, 24]. The ABC rejection algorithm takes **I**_obs_, *R*_min_, *R*_max_, *ε*, *n*_samples_ and *n*_rep_ as inputs. Here, *ε* is a tolerance parameter, such that a draw candidate *R^∗^_0_* is accepted if *RMSE < ε*; *ε* is not fixed but is auto-calibrated to keep the best 20% of all draws: *ε* = *F^−1^*_*RMSE*_(*ζ*), where *F^−1^_d_*(*ξ*) is the *ξ*-quantile of the empirical distance distribution *RMSE*_1_*, . . . , RMSE_n_*_samples_ , with *ξ* = 0.2. The rejection algorithm draws candidates *R^∗^* from the prior and performs *n*_rep_ = 20 simulations of the network SEIR model; since the network model is stochastic, the results are averaged together to produce a smooth simulated curve for **Ī**_sim_(*t*). The rejection steps are performed for candidates *R^∗^* drawn *n*_samples_ times, to densely cover the plausible range for *R*_0_; we use *n*_samples_ = 500. Once all draws are complete, the accepted values collectively form an accepted set A*_ε_*, which is an approximate sample for the true posterior *p*(*R*_0_ | **I**_obs_); note that A*_ε_* converges to *p*(*R*_0_ | **I**_obs_) if *ε* → 0. From A*_ε_*, the ABC scheme returns the median *R̂* and the credible interval CI_95%_.

During the estimation of *R*_0_, the incubation period *T_α_* is fixed to the median value obtained from epidemiological studies. Although a plausible range for *T_γ_* is available (see Table 1), no precise median value has been reported. We therefore estimate it by first applying the ABC fitting procedure to the 2018 outbreak data before control measures were introduced, fixing *R*_0_ = 2.12 as reported by [13] and fitting *T_γ_*, see Supplementary Sec. S3. This yields a median infectious period of *T_γ_* = 10 days, corresponding to *γ* = 0.1. The estimation of *R*_0_ for the 2026 outbreak then assumes that the biological characteristics of ANDV have remained unchanged over time, allowing the use of *T_α_* and *T_γ_* inferred from the 2018 outbreak.

The choice of ⟨*k*⟩ = 6 is an informed estimate, as discussed previously. However, since *R*_0_ = (*β*⟨*k*⟩)*/γ* in the small-world network model, estimating *R*_0_ effectively corresponds to estimating the product *β*⟨*k*⟩. The ABC procedure adjusts *β* for the chosen value of ⟨*k*⟩, meaning that the inferred *R*_0_ is largely insensitive to this assumption. Variations in ⟨*k*⟩ mainly affect the interpretation of *β*, provided that the network is sufficiently connected to allow the expected number of secondary infections at the start of an outbreak. In practice, ⟨*k*⟩ is required to exceed the expected range of *R*_0_, which previous studies estimate between 1 and 5 [13, 15, 24]. To support the robustness of the results for *R*_0_ and their little sensitivity to the choice of ⟨*k*⟩, we repeated the analysis with ⟨*k*⟩ = 4, 8, and 10. The resulting *R*_0_ estimates (median and IQR) differed only at the third significant digit, confirming that the inferred reproduction numbers are not sensitive to the chosen network degree. Fitting the complete 1996 and 2018 outbreaks confirmed that, although limited incidence data increase posterior uncertainty, the resulting estimates remain statistically compatible with values reported in the literature and obtained through alternative approaches.

Finally, we confirm the robustness of the results against the choice of the base network, by repeating the ABC fitting procedure on the 2026 outbreak using a scale-free network, another paradigmatic alternative to model disease spread in societies [44, 47]. To be comparable with the base small-world network, we set *N* = 150, *m* = 3 (number of edges associated with each newly added node) so that ⟨*k*⟩ ≃ 6 and the other parameters as above. The result is *R*_0_ = 3.02 [1.11, 4.90], perfectly compatible (*Z* = 0.16) with the value obtained with the small-world network.

### 4.4 Synthesis of ***R*_0_**

To combine the *R*_0_ estimates from the three outbreaks and previous studies, we performed a synthesis to obtain a single value and more constrained credible intervals, to provide a reference value for modelling and scenario analyses. Since we focus on transmission dynamics before the implementation of control measures, we included the pre-intervention *R*^(2018)^ estimate for the 2018 outbreak from [13] and the fitted pre-intervention estimate for the 2026 outbreak, *R*^(2026)^. Because these estimates were obtained from different outbreaks and modelling approaches, we first converted their reported confidence intervals into standard deviations *σ_l_*, assuming that a Gaussian approximation adequately represents the uncertainty and heterogeneity between methods, as commonly done in meta-analyses. We then combined the estimates into a single value *R̂*_0_ and standard deviation *σ̂* using the usual inverse-variance weighted formula:

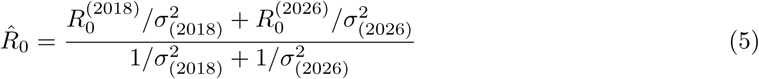

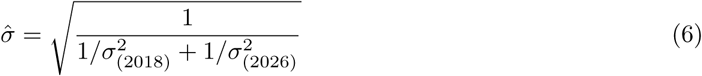

### 4.5 Model analysis, simulations and scenarios development

Results about the stability and asymptotic behaviour of the network SEIR model are known in the literature, as well as its threshold behaviour determined by *R*_0_ [36, 44, 45, 46, 47]. Nonetheless, the network model possesses degrees of uncertainties and stochasticity, linked to the network structure obtained through a rewiring probability *p*, the chance of encounters and transmission between the nodes, as well as the uncertainties associated with the parameters. We thus employ Monte Carlo Euler-Maruyama discrete-time numerical simulations to investigate the impact of parametric uncertainties and to estimate quantitative scenarios associated with plausible settings for future Hantavirus outbreaks.

#### 4.5.1 Impact of uncertainty in the parameters

To analyse the impact of uncertainty in individual parameters, such as *R*_0_ or *T_α_*, on epidemic dynamics, we perform Monte Carlo simulations using the baseline dynamical network model described above. Parameter values are sampled from their corresponding distributions (with *n* = 100 samples per simulation), while all other parameters are fixed at their median values (Table 1). Uncertainties are therefore considered one parameter at a time.

For *R*_0_, samples are drawn from a Gaussian distribution with mean *R̂*_0_ and standard deviation *σ̂* obtained from Eqs. (5) and (6) to perform an uncertainty analysis.

For *α* (and correspondingly *T_α_*), only minimum, median, and maximum values are available from the literature (see Table 1), with no information on the underlying distribution. Following recent studies suggesting that Gamma distributions provide a suitable approximation for epidemiological parameters [48, 69], we estimate a Gamma distribution Γ(*α^′^, θ^′^*) by least-square fitting its quantiles to the available values. Specifically, we constrain the 95% credible interval between the minimum incubation period (*T*^(*min*)^_α_ = 7 days) and maximum value (*T*^(*max*)^_α_ = 42 days), with median *T̂*_α_ = 18 days. The resulting parameters are *α^′^* = 4.74 (scale parameter) and *θ^′^* = 4.24 (shape parameter), giving quantiles *Q*(2.5%) = 6.27, *Q*(50%) = 18.69, and *Q*(97.5%) = 41.80 days. The corresponding mean is *T̃*_α_ = 20.1 days, indicating moderate right skewness relative to the median. A comparison with alternative distributions (a Poisson with the same mean and a Gaussian with the same mean and standard deviation set to have *Q*(97.5%) = 42) is reported in Supplementary Fig. S4. Monte Carlo samples are therefore drawn from Γ(4.74, 4.24).

On *T_α_*, we then perform two complementary analyses. First, in the uncertainty analysis, an incubation period is sampled from Γ(4.74, 4.24) at each simulation step for infected nodes, representing uncertainty in the true incubation time. This addresses the question: what does the epidemic look like when the incubation period is uncertain? Additionally, we perform a sensitivity analysis: a single value of *T̃*_α_ is sampled from the same distribution at the beginning of each simulation and kept fixed throughout the outbreak. Here, randomness arises only from the stochastic network dynamics, addressing how epidemic outcomes vary across the plausible range of incubation periods. Results of the sensitivity analysis, its comparison with the uncertainty analysis, and a comparison with the uncertainty analysis on *R*_0_ that supports the fitting results, are reported in Supplementary Sec. S4.

#### 4.5.2 Superspreading

When superspreading (the possibility that an infectious person infects many more people than the average) is enabled, the homogeneous *β* is replaced by a node-and day-specific draw, making the transmission probability possibly node-specific and time-dependent, so as to model significant heterogeneity. We first use the Lloyd-Smith et al. [70] parametrization for superspreading, generalising the Negative Binomial distribution in Supplementary Sec. S1 with a Gamma distribution Γ*_ν_* (*k_d_, R*_0_*/k_d_*), as done for other parameters; *k_d_* → ∞ corresponds to the homogeneous case. The default *k_d_* is set to 0.394, from the fit in in Supplementary Sec. S1. We then perform a sensitivity analysis around it. For node *i* on day *t*, we sample *ν_i_*(*t*) from Γ*_ν_* and set the transmission probability as

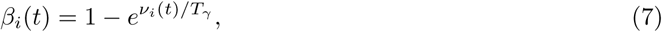

so that *β_i_*(*t*) is used in the Bernoulli trial to infect each neighbour. The formulation returns the negative binomial offspring distribution introduced in [70] to model superspreading. The Gamma parametrization preserves E(*β_i_*) = *β* and E(*ν*) = *R*_0_; only the variance of new infections increases as the dispersion parameter *k*_d_ decreases, as *V ar*[*ν*] = *R*^2^_0_/*k_d_*.

A node is classified as a superspreader on a given day when it causes more than 4 offsprings among its neighbours. The threshold is chosen so that it exceeds *R̂*_0_ for more than 3*σ̂*. The count of superspreading events cumulate all events with superspreader nodes within a Monte Carlo simulation, performed on the network model as described above..

#### 4.5.3 Control strategies

For scenarios where control interventions are enacted after time *t*_int_, we augment the set of system’s states with *T* (Traced, removed from the Exposed pool with a constant tracing rate *χ*) and *Q* (Quarantined, removed from the Infectious pool with rate *η*). In this case, *x^′^*(*t*) ∈ {*S, E, I, R, T, Q*}, and the flow diagram includes the orange flows in Fig. 2; the conservation law *S* + *E* + *I* + *R* + *T* + *Q* = *N* holds at each time step. For the transition out of *x_i_*(*t*) = *E*, for *t < t*_int_ the governing rule is the standard *P* (*x_i_*(*t* + 1) = *I*|*x_i_*(*t*) = *E*) = *α*. For *t* ≥ *t*_int_, an exposed node faces two competing daily hazards: progression to infectiousness, with the same rate *α*, or isolation with a rate *χ*. In the implementation, these are resolved via sequential Bernoulli trials, checking isolation first, so that *P* (*x_i_*(*t* +1) = *T* |*x_i_*(*t*) = *E*) = *χ* and *P* (*x_i_*(*t* +1) = *I*|*x_i_*(*t*) = *E*) = (1 −*χ*)*α*. Similarly, for the transition out of state *x_i_*(*t*) = *I*, the possibility of being quarantined is prioritized against the possibility of being removed when *t* ≥ *t*_int_: *P* (*x_i_*(*t*+1) = *Q*|*x_i_*(*t*) = *I*) = *η* and *P* (*x_i_*(*t*+1) = *R*|*x_i_*(*t*) = *I*) = (1−*η*)*γ*. Note that quarantine takes effect for the following day’s transmission opportunities, under the assumption that detection happens after exposure has already occurred during that day. Traced and Quarantined nodes are assumed to not contribute anymore to the infection dynamics, as a minimum of 42 days of quarantine was always enforced [27]; hence, nodes from *x_i_*(*t*) = *T* and *x_i_*(*t*) = *Q* directly transition to state *x_i_*(*t*) = *R* with the same rate *γ*.

#### 4.5.4 Asymptomatic cases

For scenarios accounting for the possibility of pre-symptomatic infectiousness, we augment the set of system’s states with *A* (Asymptomatic), so that *x^′′^* = {*S, E, I, A, R, Q*}. We keep the *Q* state and its associated transition rates to investigate the effect of quarantining only symptomatic individuals *I*; *A* are never quarantined. The state *T* is not considered, as testing and isolation only applies to exposed individuals; the additional complexity of asymptomatic transmission reducing the effectiveness of contact tracing is not considered. The flow diagram for the SEIARQ model is shown in Supplementary Sec. S6. At each time *t*, only a fraction *θ* of individuals in *E* move to *I* (symptomatic infectious), while a fraction (1 − *θ*) transitions to *A*. Individuals in *A* can then move to *I* with a rate *ζ* = 1*/T_ζ_* or directly to *R* at a rate *γ*; since the model allows for both routes (individuals can be infectious before and after developing symptoms, or never become symptomatic), we use “pre-symptomatic” and “asymptomatic” infectiousness as synonyms. To maintain a causal flow between infectious compartments, we pose that the transition between *E* and *A* occurs with a probability *α*_1_ = *α* · *a*, with *a >* 1: it is faster than the transition *E* → *I*. Consequently, *T_ζ_* is constrained such that *T_α_* = *T_α_/a* + *T_ζ_*. This way, we maintain the interpretation and values of the incubation rate. Studying the sensitivity to *T_ζ_* implies studying the sensitivity to *a*. In general, symptomatic and pre-symptomatic individuals may infect with different probabilities *β* and *β_A_*, respectively. We here assume *β_A_* = *β*; alternative probabilities should be set after detection and assessment of asymptomatic transmission and are left for future studies. *R*_0_ is thus unchanged. In the implementation, the branching probabilities across states are solved with Bernoulli trials: from *E*, the asymptomatic branch is checked first; from *A*, we first check progression to *I*. For the transitions between the other compartments, we use the same rules and probabilities as before.

## Data and code availability

The database consisting of incidence and timing for the 1996, 2018 and 2026 outbreaks is accessible at the public repository https://github.com/daniele-proverbio/hantavirus, together with the code used for the analysis and for the simulations.

## Acknowledgments

We thank Cristina Mussini e Andrea Cossarizza for useful discussions about the early stages of the 2026 outbreak and its clinical aspects.

## Author contributions

D.P.: Conceptualization, Methodology, Software, Validation, Formal analysis, Investigation, Data curation, Visualization, Writing - original draft, Writing - review and editing. G.G.: Conceptualization, Writing - review and editing, Funding acquisition, Supervision, Project administration.

