## Supplementary Material for "*Andes* Hantavirus human-to-human transmission dynamics and preparedness scenarios"

#### S1 Statistics of past outbreaks and test for superspreading in ANDV outbreaks

Assessing superspreading in ANDV transmission requires reconstructing transmission chains, estimating the number of secondary infections $Z$, and identifying the statistical distribution that best describes it [1]. Early epidemic spread can be approximated by a branching process [2], where $Z$ follows a Poisson distribution with mean $R_0$ [3]. To account for individual heterogeneity, epidemiological models instead assume a gamma-distributed infection potential, yielding a Negative Binomial distribution, $Z \sim \text{NegBinom}(R_0, k_d)$, which is argued to more effectively describe epidemics where superspreading is a key feature [1, 4]. Here, $R_0$ is the average value, while $k_d$ is the dispersion parameter: smaller values indicate greater heterogeneity and a higher probability of superspreading. The variance is given by $R_0(1 + R_0/k_d)$ and, depending on $k_d$, the distribution can be more or less skewed to the right, thereby capturing superspreading events with higher probability than a simple Poisson process. The Negative Binomial and Poisson models can be compared using the Akaike Information Criterion (AIC) [5] after fitting them to data. The fitting procedure not only tests whether superspreading may better describes the transmission tree of an outbreak, but it also returns the parameters for a Poisson or Negative Binomial distribution, which can be used to quantitatively inform the construction of realistic scenarios.

The 1996 outbreak showed no evidence of superspreading [6]; accordingly, the Poisson model provided a slightly better fit than the Negative Binomial model [1, Tab. 1] ($\text{AIC}_{\text{Poisson}} < \text{AIC}_{\text{NegBinom}}$). For the 2026 ship outbreak, transmission chains or information about $Z$ are unavailable, preventing this analysis.

In contrast, the 2018 outbreak was associated with superspreading, driven by symptomatic individuals attending crowded events such as a birthday party and a wake [7]. Our analysis (Fig. S1) confirms that the Negative Binomial distribution provides a better fit than the Poisson model ($\text{AICPoisson} = -2.8 > \text{AICNegBinom} = -4.8$). The estimated parameters are $R_0 = 2.77$, consistent with the pre-intervention estimate of 2.12 (95% CI: 1.24–3.35) reported in [7], and $k_d = 0.394$, comparable to estimates for COVID-19 outbreaks worldwide of $k_d^{(\text{COVID})} = 0.41[0.23; 0.60]$ (mean and IQR) [8], with extremes at 0.04 [9] and 2.97 [10]. $R_0 = 2.77$ and $k_d = 0.394$ correspond to a variance of 5.79 and a number of successful infection $n = 2.536$ with probability $p = 0.478$. and define the Negative Binomial parameters used to construct epidemic scenarios that explicitly account for superspreading. These values are used to quantitatively inform the scenarios in Main Text where superspreading is explicitly taken into account.

#### S2 The Watts-Strogatz model

The contact network is constructed via the standard procedure for a Watts-Strogatz model [11], see Fig. S2. $N$ nodes are placed on a $k$-regular ring lattice, connecting node $i$ (e.g., the circlced one) to its $k/2$ nearest neighbours on each side (Fig. S2, left); in the example, $k = 4$. For each edge $(i, j)$, we independently rewire $j$ to a uniformly random node $j' \notin \{i\} \cup \mathcal{N}'(i)$ with probability $p$, where $\mathcal{N}'(i) \subseteq \mathcal{N}$ is the adjacency set of node $i$: in Fig. S2, centre, the rewired edges are in orange. We do this for all nodes, using by default $p = 0.1$, which is a typical value for a small-world network [12]. The

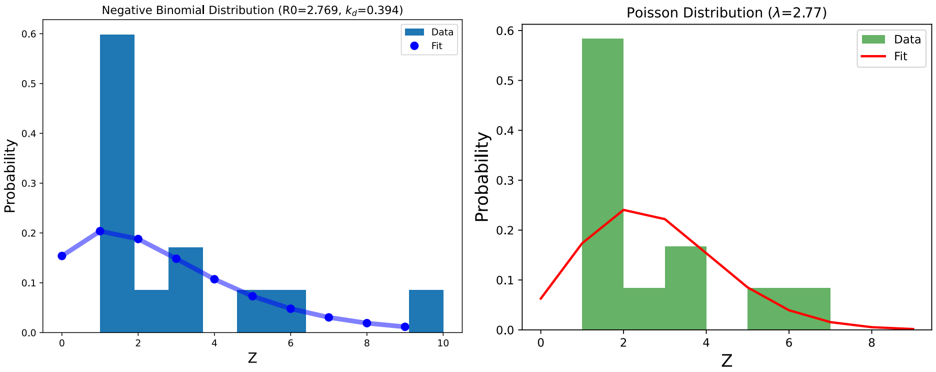


Figure S1: **Fitting overdispersion distributions to data.** Left: fitting a Negative Binomial distribution to data about offsprings $Z$ from the 2018 outbreak. Right: fitting the same data using a Poisson distribution.

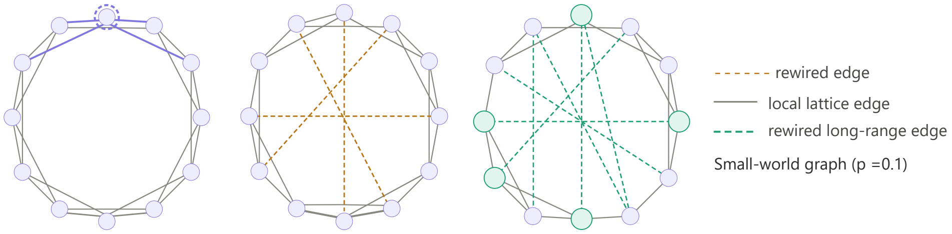


Figure S2: **Construction of the Watts-Strogatz model.** Left: a ring lattice with $N$ nodes, with mean degree $k$. Centre: the begin of the rewiring process. Left: the final network, with short and long-range edges.

process creates a network with local (in grey) as well as long-range (green) edges, see Fig. S2, right. The mean degree $\langle k \rangle$ is thus preserved by the rewiring procedure and is equal to $k$.

### S3 Estimation of the infectious period

The infectious period $T_\gamma$ is reported to lie between 7 and 14 days [13, 14], but no reliable median estimate is available. We therefore infer $T_\gamma$ from data collected during the 2018 outbreak, using independent estimates of $R_0$ to obtain reliable values for $T_\gamma$ and calibrate plausible scenarios.

Using pre-intervention data (recall Fig. of Main Text), we fix the incubation period to $T_\alpha = 18$ days and set $R_0 = 2.12$ [7]. Applying the ABC fitting procedure (as described in the Methods) with a uniform prior, $T_\gamma \sim U(7, 14)$, yields $\hat{T}_\gamma = 9.86$ $[7.22, 13.33]$ days (median and IQR).

We improve the reliability of this estimate with three independent tests. First, we repeat the analysis using $R_0 = 2.77$, obtained independently in Sec. S1. We obtain the same result, $\hat{T}\gamma = 9.86$ $[7.22, 13.48]$ days, confirming robustness to the choice of $R_0$.

Second, we modify the ABC error by using a composite distance that also imposes a constraint on the serial interval. The serial interval SI (time between successive cases in a chain of transmission) was estimated in [7] to be $23 \pm 7$ days; since it corresponds, approximately, to $\mathrm{SI} = T_\alpha + 2T_\gamma$, it can be used to add a constraint to the ABC procedure, via inclusion in the distance metric as

$$d_{\text{total}} = (1 - w) \cdot RMSE + w \cdot d_{\text{SI}} \ , \tag{S1}$$

where $RMSE$ is the same distance between observed data and the simulated curve, as in Eq. 3 of Main Text and $d_{\text{SI}} = |\mathrm{SI}_{\text{sim}} - \mathrm{SI}_{\text{obs}}| / \mathrm{SI}_{\text{sim}}$ is the relative error in the estimation of the SI. $w$ acts as a weight on the two terms of the distance metric: we use $w = 0.7$ to give more weight on the deviations from the observed data. The result is $T_\gamma = 9.98$ $[9.27, 10.78]$ days, consistent with the values obtained previously. The estimated SI is of 22.93 days, very close to SI= 23 days estimated in literature.

Finally, we run a fit for $R_0$ using the very same data employed to estimate $T_\gamma$, which is now held fixed. This way, we cross-check that the epidemiological parameters are properly calibrated.

This re-estimation gives $R_0 = 1.23$ $[0.63, 2.19]$, which is statistically consistent with the value of $R_0 = 2.12$ $[1.24, 3.35]$ from [7]: A Z-test between the two estimates gives $Z_{\text{score}} = 1.17$, which does not reject the null hypothesis of compatibility.

All subsequent analysis thus uses $T_\gamma = 10$ $[9, 11]$, after rounding the results to the smallest measurement unit of one day.

### S4 Focus on the incubation period

#### S4.1 Gamma distribution: inference and interpretation

As described in Methods, we have inferred the distribution of the incubation period $T_\alpha$ by fitting a constraint Gamma distribution $\Gamma(\alpha', \theta')$ to the minimum, median and maximum values provided in previous work [15]. The shape of the estimated Gamma distribution, compared with a Poisson distribution with the same mean and a Gaussian distribution with the same mean and std such that $Q(97.5\%) = 42$, is shown in Fig. S3.

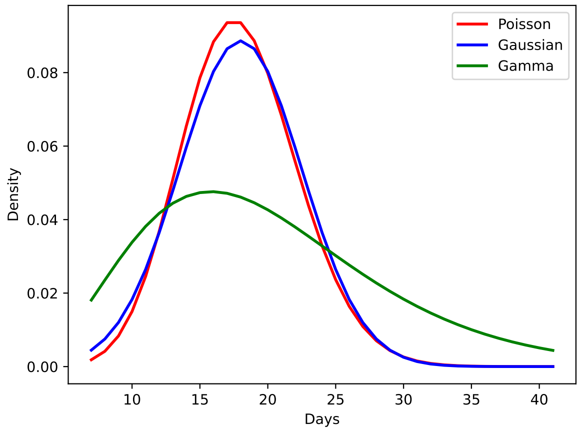


Figure S3: **Comparison of Gamma, Poisson and Gaussian distributions to model uncertainties on $T_\alpha$.** In green, the $\Gamma(4.74, 4.24)$ distribution obtained from fitting the data from literature. In red, a Poisson distribution with mean equal to 20.1 (mean of the Gamma distribution). In blue, a Gaussian distribution with the same mean; the standard deviation is of 4.5 days, so that $Q(97.5\%) = 42$.

#### S4.2 Sensitivity analysis

As described in the Main Text and Methods, we perform a sensitivity analysis on $T_\alpha$ in addition to the uncertainty analysis. While the two approaches capture different sources of stochasticity (network structure alone versus network structure combined with uncertainty in the incubation period), they produce similar ensemble outcomes. As shown in Fig. S4, individual trajectories differ from those in Fig. 4 of the Main Text, but the overall statistics remain comparable.

The visual result is confirmed by a Kolmogorov–Smirnov test, which shows no significant differences (p-value $> 0.05$) in the distributions of peak height, peak time, or attack rate between the sensitivity and uncertainty analyses (compare with Fig. 4 of Main Text), or between the uncertainty analyses for $T_\alpha$ and $R_0$ (compare with Fig. 3, Main Text). These results indicate that scenario predictions can focus on uncertainty in $R_0$, as its estimation already captures the effects of uncertainty in $T_\alpha$.

#### S4.3 Delay SEIR model

The main ANDV analysis uses the network model described in the Main Text. To complement it with a framework that enables formal analysis and control, we also consider a well-mixed delay ODE SEIR model like in [16]. Unlike the network model, this approach neglects contact heterogeneity but allows us to isolate the effects of uncertainty in the incubation period $T_\alpha$, governing the transition from the $E$ to $I$ compartments. At the start of each Monte Carlo simulation, $T_\alpha$ is sampled from the same

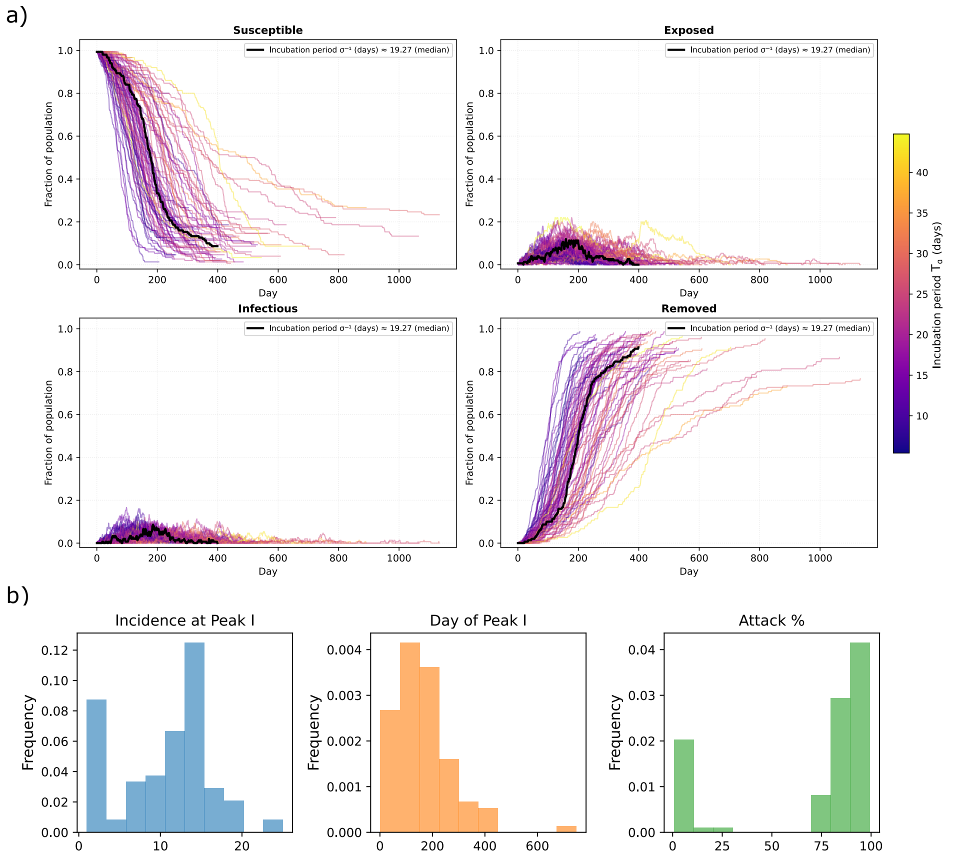


Figure S4: **Sensitivity analysis on $T_\alpha$.** a) Trajectories of the $S$, $E$, $I$ and $R$ states, obtained with Monte Carlo simulations of the network model; for each simulation, $T_\alpha$ is drawn once from a Gamma distribution with 95% CI in $[7, 42]$ (see details in Methods). b) Statistics of the ensemble of trajectories: maximum incidence at peak, timing of the peak, and total attack rate.

Gamma distribution used in the network analysis, $\Gamma(4.74, 4.24)$, with mean $\simeq$ 20 days, and remains fixed throughout that run.

To represent an uncertain incubation period, the exposed compartment $E$ is divided into $m$ sequential sub-compartments $E_1, \ldots, E_m$, each representing one identically-paced stage of the latent period. Newly exposed individuals progress through all stages before becoming infectious. We set $m = \text{round}(\alpha')$, so the resulting Erlang distribution closely approximates the prior Gamma distribution. For each ODE simulation, the transition rate is $\lambda_r = m/T_\alpha^{(r)}$, yielding a deterministic trajectory conditional on the sampled incubation period. The total time spent crossing all stages is the sojourn time in $E$, which follows an Erlang distribution $Er(m, \lambda_r)$; this is a special case of the Gamma distribution and is widely used to represent disease stages in compartment models [17, 18].

The transmission rate $\beta$ is chosen to match the ANDV $R_0$ of the network model, allowing direct comparison; the linear chain does not alter $R_0$ because the exposed sub-stages do not transmit. The system is integrated with Python's RK45 solver over $\mathcal{T} = 500$ days; repeating the process for multiple draws produces an ensemble of epidemic trajectories reflecting uncertainty in $T_\alpha$.

Figure S5 shows 100 such trajectories, coloured by the sampled incubation period. Shorter incubation periods produce earlier, sharper epidemic peaks, whereas longer periods delay and flatten the peak. As expected, uncertainty in $T_\alpha$ affects the timing of the peak but not the final attack rate, which is determined by $R_0$. The quantity reported as $E(t)$ in the output is the sum across all sub-stages:

$$E(t) = \sum_{j=1}^{m} E_j(t) \tag{S2}$$

which represents the total number of exposed (incubating) individuals at time $t$.

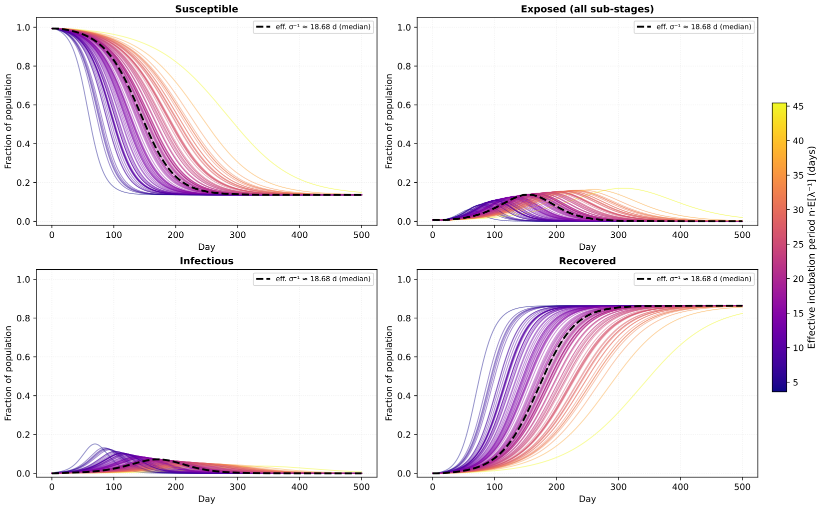


Figure S5: **Uncertainty analysis using the delay SEIR compartment model.** Shown, are the trajectories of the $S$, $E$, $I$ and $R$ states.

Finally, we perform a sensitivity analysis on the delay SEIR model by varying $T_\alpha$ deterministically across simulations. As shown in Fig. S6a, the resulting trajectories closely match those obtained from the uncertainty analysis (Fig. S5), as variability is smoothed by the delay chain. Figure S6b reports the distributions of peak height and timing, which are qualitatively similar to those obtained with the network model. However, a Kolmogorov–Smirnov test yields p-value $< 0.05$, indicating statistically significant differences between the two approaches. This is expected, as the ODE delay SEIR model represents the limiting case of a highly connected network, with $p_r = 1$ and large $\langle k \rangle$. Overall, the results remain consistent at a qualitative level, suggesting that deterministic delay-based models can effectively complement network models in the study of ANDV epidemics.

### S5 Sensitivity on network structure

As described in the Main Text, we investigate the role of the network structure by varying the network parameters $\langle k \rangle$ and $p_r$. While Fig. 5 of the Main Text reports ensemble results, here we present representative single trajectories for each parameter value. Figures S7, S8, and S9 show sensitivity analyses varying one parameter at a time while keeping the others fixed to the default values in Table 1 of the Main Text.

Because the network model is stochastic, individual epidemic trajectories are not necessarily ordered by increasing $\langle k \rangle$ or $p_r$ and may show random deviations. Comparing Figs. S7 and S8 confirms that larger networks (with higher $N$) produce smoother trajectories while preserving the overall epidemic dynamics. In Fig. S9, the most distinct trajectory corresponds to $p_r = 0$, representing a purely lattice network. This case is shown for completeness but is unrealistic and excluded from the other analyses.

### S6 Asymptomatic infectious carriers

The possibility of pre-symptomatic transmission [19] motivates further analysis of the effectiveness of quarantining infectious individuals. Figure S10 shows the SEIARQ model used in the Main Text, implemented on the same network framework. The model introduces an asymptomatic infectious state $A$: a fraction $\theta$ of exposed individuals become symptomatic at rate $\alpha$, while the remaining fraction $(1 - \theta)$ enters $A$. Asymptomatic individuals develop symptoms after $T_\zeta > 0$ days or recover directly.

Here, we focus on quarantine at rate $\eta$ as the main control strategy. Testing and isolation at rate $\chi$ only applies to exposed individuals; although asymptomatic transmission may reduce the effectiveness of contact tracing, this additional complexity is not considered and is instead assumed to be reflected by a lower effective $\chi$. For the interpretation of states and rates, refer to the Main Text.

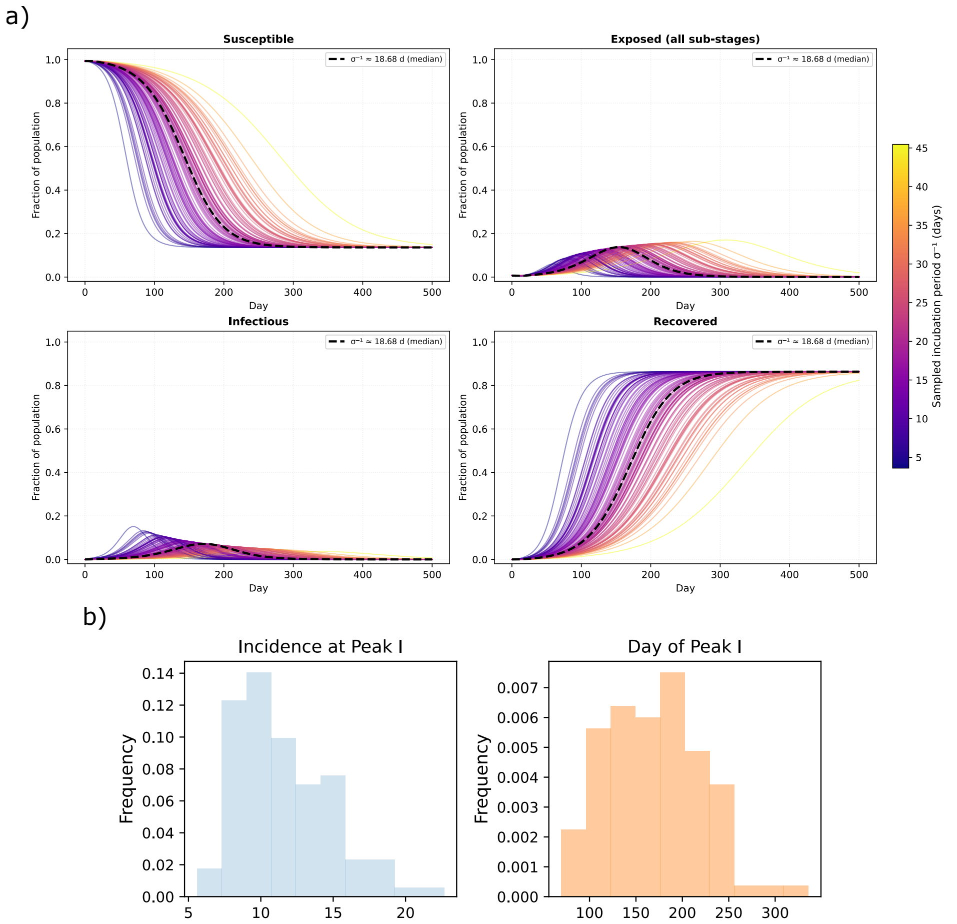


Figure S6: **Uncertainty analysis using the delay SEIR compartment model.** a) The trajectories of the $S$, $E$, $I$ and $R$ states. b) The height and timing of the peak. The attack rate is not shown as all curves converge to the disease-free equilibrium within $\mathcal{T} = 500$ days.

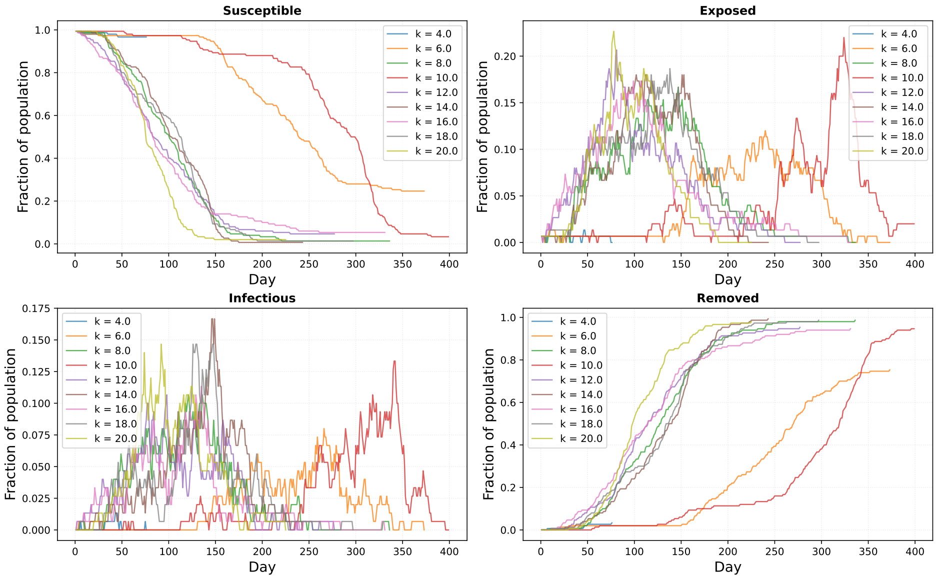


Figure S7: **Sensitivity analysis on $\langle k \rangle$, on a network with $N = 150$ nodes**. Shown, are examples of single trajectories of the $S$, $E$, $I$ and $R$ states.

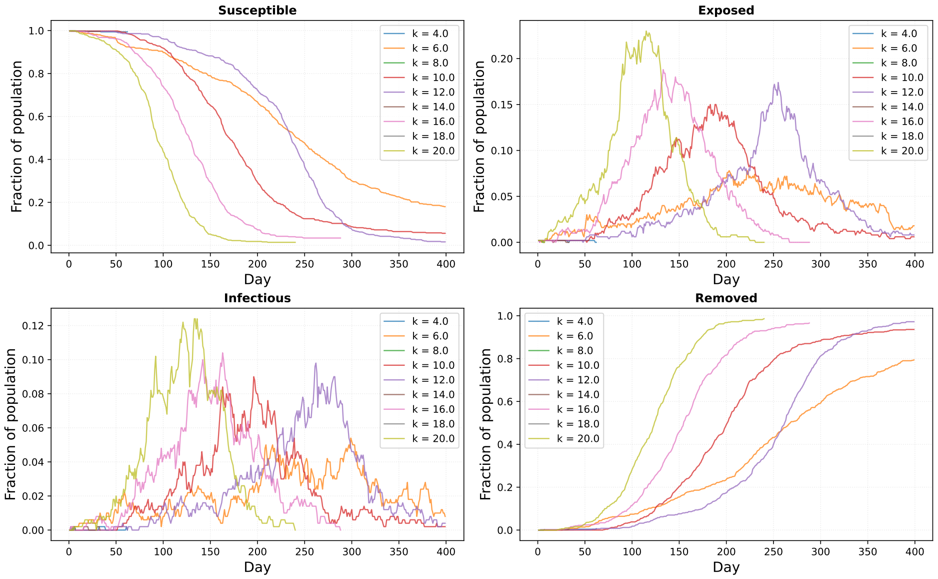


Figure S8: **Sensitivity analysis on $\langle k \rangle$, on a network with $N = 500$ nodes.** Shown, are examples of single trajectories of the $S$, $E$, $I$ and $R$ states.

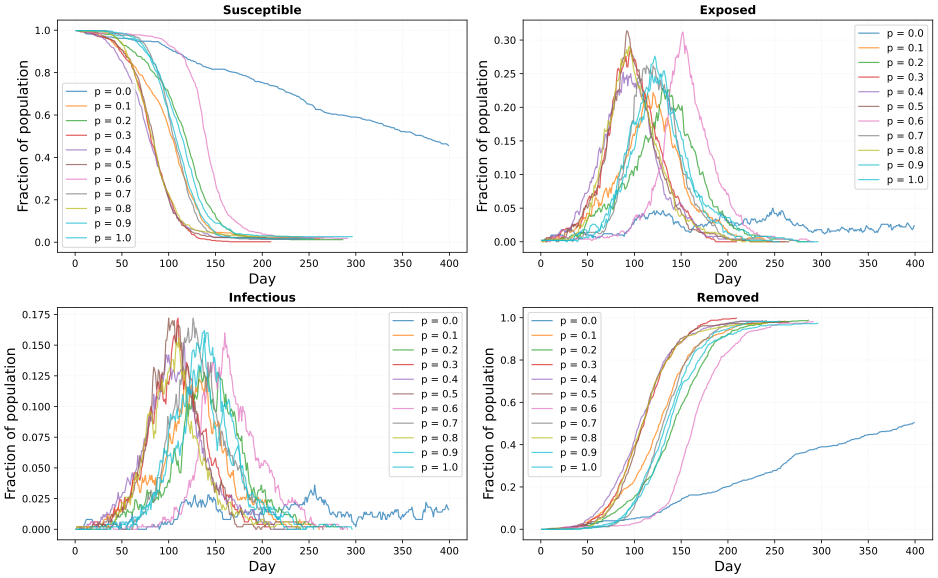


Figure S9: **Sensitivity analysis on $p_r$, on a network with $N = 150$ nodes.** Shown, are examples of single trajectories of the $S$, $E$, $I$ and $R$ states.

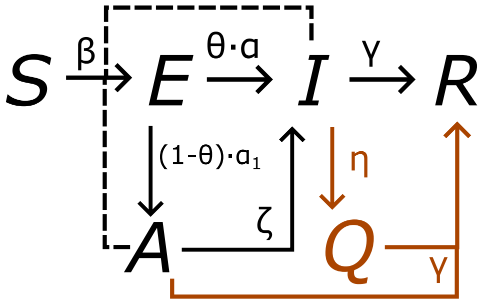


Figure S10: **Model scheme of the epidemic SEIARQ model, with quarantine of infectious symptomatic individuals.** The new state $A$ corresponds to asymptomatic infectious individuals.
